# Protein risk scores enable precise prediction of cardiovascular events in chronic kidney disease patients

**DOI:** 10.1101/2025.07.25.25332196

**Authors:** Yang-Gyun Kim, Yonghyun Nam, Thomas M Westbrook, Jaehyun Joo, Jakob Woerner, Rajat Deo, Marylyn D Ritchie, Dokyoon Kim

## Abstract

**Background:** Cardiovascular disease (CVD) is the leading cause of death in patients with chronic kidney disease (CKD). However, there is still a lack of reliable biomarkers to predict cardiovascular events (CVEs) in this population.

**Methods:** This study aimed to develop a protein risk score (ProRS) model to predict CVEs in CKD patients. From the UK Biobank Pharma Proteomics Project (UKB-PPP), a total of 1,799 patients with CKD and no prior history of CVD were enrolled. Participants were randomly divided into a training set (70%) and an evaluation set (30%). We analyzed 2,920 plasma proteins to identify associations with CVEs, including coronary heart disease, heart failure, and ischemic stroke.

**Results:** After adjusting for significant clinical factors, 38 proteins remained consistently significant in both the training and evaluation sets. Using an elastic net model, we selected 34 to construct the ProRS. The area under the receiver operating characteristics curve for annual CVEs prediction using the ProRS ranged from 0.67 to 0.74, compared to 0.60 to 0.69 for a clinical risk model, and 0.58 to 0.63 for a polygenic risk score. The 10-year incidence of CVEs among individuals in the top 5% of the ProRS distribution was 44.4%, significantly higher than 29.6% observed in the top 5% of the clinical risk model. Conversely, the bottom 5% of the ProRS group showed a 0% incidence rate, compared to 3.7% in the bottom 5% of the clinical risk model, demonstrating superior performance in both risk identification and exclusion. Notably, among patients classified as low risk by the clinical risk model, those with a high ProRS showed an increased risk of CVEs. In contrast, when the ProRS was low, the influence of the clinical risk model on event prediction was minimal. Mendelian randomization analysis identified 25 proteins whose levels were causally influenced by CKD, 10 of which were also associated with CVD.

**Conclusions:** We demonstrated that plasma proteomics holds promise as a predictive biomarker for CVEs in patients with CKD. By enabling early identification of high-risk individuals, this approach may facilitate timely preventive interventions and ultimately reduce cardiovascular mortality in this vulnerable population.

## 1. Introduction

Chronic kidney disease (CKD), affecting 12.3% of adult worldwide, is a progressive condition with no specific cure, ultimately leading to end-stage renal disease (ESRD), where patients can survive only with renal replacement therapies ^1^. Cardiovascular disease (CVD) remains the leading cause of morbidity and mortality among individuals with CKD, accounting for more than half of all deaths in this population ^2^. In particular, the cardiovascular mortality in ESRD patients is 10 to 20 times higher than in healthy individuals ^3^. The link between CKD and CVD is multifactorial, involving hemodynamic overload, systemic inflammation, oxidative stress, and shared metabolic risk factors ^4,5^. This is largely due to the cardiorenal syndrome, a pathological interaction between the heart and kidney that mutually exacerbates dysfunction, and the fact that certain cardiovascular treatments are limited in CKD patients ^6,7^.

However, there are no clinical guidelines or biomarkers available for the primary prevention of CVD in CKD. Although a decline in estimated glomerular filtration rate (eGFR) is considered one of the most significant risk factors for CVD, there are no specific cholesterol management guidelines based on kidney function and proteinuria. In addition, clinical risk prediction tools, such as the Pooled Cohort Equations (PCE), which rely largely on traditional risk factors, provide limited insight into the molecular processes driving CVD in the setting of CKD ^2^. Currently used biomarkers, such as troponin and B-type natriuretic peptide, have limited predictive value for CVD due to altered metabolism or excretion in CKD ^8^. While polygenic risk scores (PRS) have been used to predict complex diseases, they often lack mechanistic specificity and may not fully capture disease-relevant biology in high-risk subgroups such as CKD patients ^9^. There is a growing interest in identifying circulating biomarkers that capture early and mechanistically informative signals of cardiovascular risk. Among these, plasma proteins have emerged as promising candidates. Recently, proteomic studies from the Chronic Renal Insufficiency Cohort (CRIC) and Atherosclerosis Risk in Communities (ARIC) cohorts have identified proteins associated with incident CVD in patients with CKD ^10^. Nevertheless, obtaining large-scale high-throughput proteomics data that aligns with specific research objectives remains a significant challenge. As a result, few studies have applied this approach to cardiovascular risk prediction in the context of CKD.

In this study, we aimed to identify plasma proteins associated with incident cardiovascular events (CVEs) among individuals with CKD using UK Biobank Pharma Proteomics (UKB-PPP). Prevalent CKD patients were selected from the UKB-PPP cohort and randomly assigned to either a discovery set (70%) or an evaluation set (30%). First, we sought to identify proteins significantly associated with incident CVD in patients with CKD. Second, we aimed to develop a protein risk score (ProRS) to predict CVD occurrence in this population. Third, we explored the biological significance of these proteins in the context of CVD development in CKD. Finally, we compared the predictive performance of the ProRS with that of conventional cardiovascular risk models.

## 2. Methods

### 2.1 Study design and cohorts

This study utilized plasma proteomics data from the UK Biobank (UKB), which is linked to genetic data and electronic health records. Among ∼500,000 UKB participants, plasma proteomic measurements were available for 44,194 individuals enrolled in the UKB Pharma Proteomics Project (UKB-PPP), comprising 2,923 proteins quantified using the Olink Explore 3072 platform ^11^. We identified 1,799 individuals with prevalent CKD and no history of CVEs at baseline for analysis. CKD was defined as the presence of CKD-related diagnostic codes or an eGFR < 60 mL/min/1.73 m^2^ at baseline. The eGFR was calculated with the CKD-EPI 2021 equation based on serum creatinine (Cr) levels ^12^. Participants with ESRD, prior kidney transplantation, or any documented CVEs before baseline were excluded.

The primary outcome was incident CVE, defined as a composite of coronary heart disease (CHD), ischemic stroke (ISS), heart failure (HF), or cardiovascular mortality, based on harmonized ICD codes from linked hospital and mortality records (**Supplementary Table 1)**. Baseline was defined as the date of blood collection. Missing binary covariates were imputed using the mode, and missing continuous covariates were imputed using cohort-specific medians. Participants were followed from baseline until the occurrence of a CVE, death from non-cardiovascular causes, loss to follow-up, or administrative censoring.

For proteomics data, after quality control, normalized protein expression (NPX) values were retained for 2,920 proteins. Missing NPX values were imputed using 10-nearest neighbor imputation based on Euclidean distance in the protein expression space. Time-to-event analyses were conducted using Cox proportional hazards models. Model performance was evaluated using the concordance index (C-index) and time-dependent area under the receiver operating characteristic curve (AUC) over a 1-to 10-year follow-up period. **Figure 1** provides an overview of the study design, including (a) identification of baseline CKD patients and follow-up for incident CVEs, (b) a multi-stage framework for identifying CVE-associated plasma proteins, and (c) development and evaluation of a ProRS, benchmarked against established clinical models.

**Figure 1.**
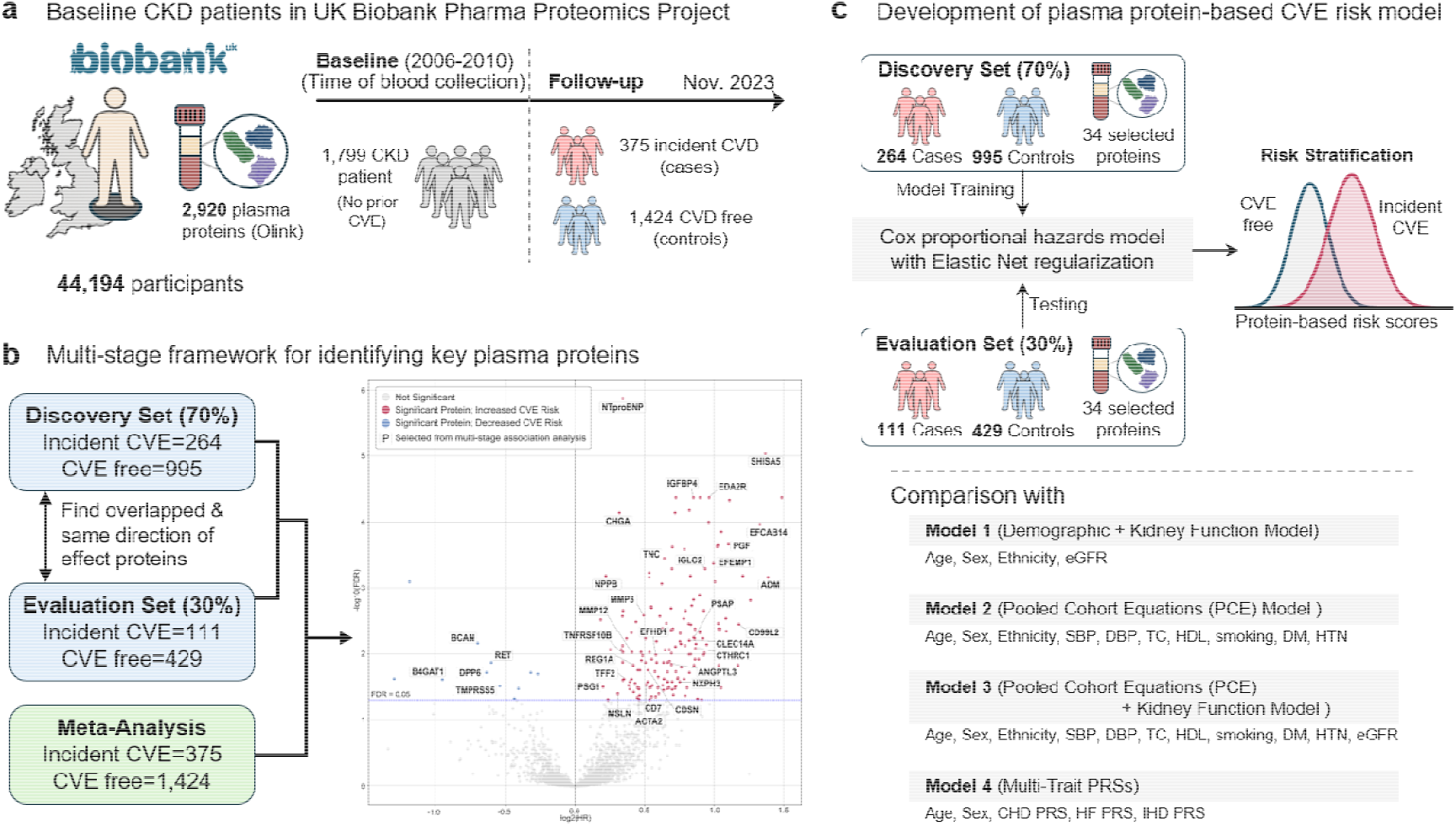
Study design, biomarker discovery, and proteomics-based risk prediction framework. (a) Overview of the study population: 1,799 individuals with CKD and no prior CVEs were selected from the UK Biobank Pharma Proteomics Project (UKB-PPP), with 2,920 plasma proteins (b) A multi-stage association analysis was performed using a 70/30 discovery-evaluation split to identify proteins associated with incident CVEs; 34 proteins meeting FDR significance and consistent effect directions were prioritized. The volcano plot displays the meta-analysis results: each point represents a protein, with the x-axis showing the log □ HR and the y-axis indicating the –log □ □ FDR-adjusted P-value. Red and blue dots highlight proteins significantly associated with increased or decreased CVE risk, respectively (q < 0.05). Proteins selected from the multi-stage analysis (n = 34) are annotated with protein name (c) A Cox model with Elastic Net regularization was trained using these proteins to derive a CVE risk score, with evaluation against four benchmark models.

### 2.2 Protein biomarker discovery using measured proteomics

To identify plasma proteins associated with incident CVEs, we applied a multi-stage Cox regression framework within the CKD cohort ^13,14^. The cohort was randomly split into a discovery set (70%) and an evaluation set (30%), stratified by age, sex, eGFR, and CVE status to preserve distributional balance. Within each subset, Cox proportional hazards models were fit separately for each protein, adjusting for age, sex, race/ethnicity (White vs. non-White), eGFR, systolic and diastolic blood pressure (SBP, DBP), hypertension (HTN), body mass index (BMI), total cholesterol (TC), high-density lipoprotein cholesterol (HDL), diabetes (DM), smoking status, and alcohol use. Proteins meeting nominal significance (P < 0.05) and showing consistent effect directions across both discovery and evaluation sets were retained. These overlapping proteins were then re-evaluated in the full cohort using the Cox proportional hazards models. Proteins passing a false discovery rate (FDR) threshold of q < 0.05 in the full analysis were selected, resulting in 34 proteins prioritized for downstream modeling (**Figure 2b**).

**Figure 2.**
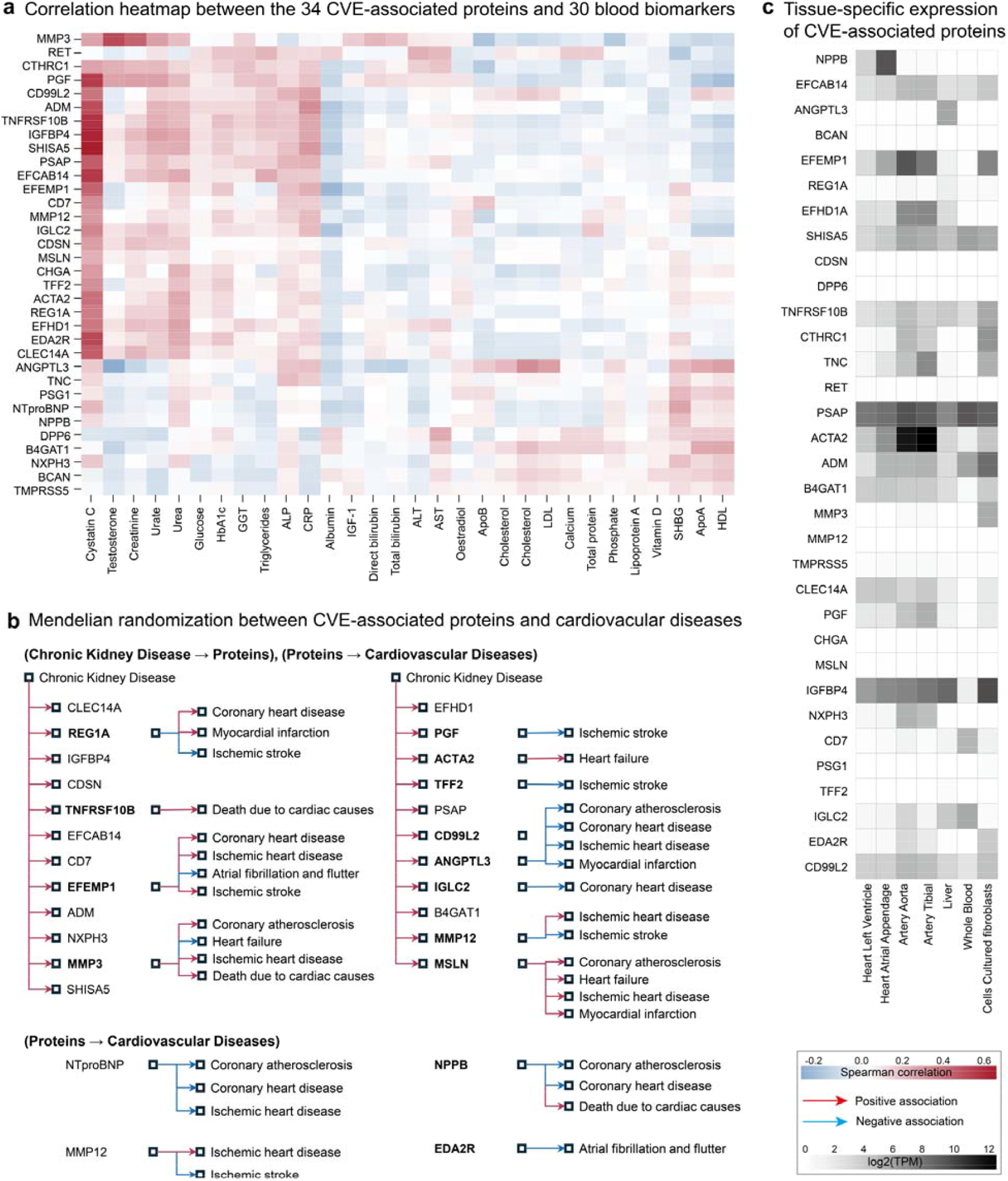
Biological characterization of CVE-associated proteins. (a) Correlation heatmap between the 34 CVE-associated proteins and 30 standard clinical blood biomarkers in the UKB-PPP cohort, with red and blue indicating positive and negative associations, respectively (P < 0.001). (b) Bidirectional Mendelian randomization (MR) analysis linking CKD to protein levels and protein levels to cardiovascular outcomes .(c) Tissue-specific expression profiles of the 34 proteins based on GTEx RNA-seq data, with each protein assigned to its highest-expressing tissue.

### 2.3 Biological Meaning of Associated Proteins

To investigate the biological relevance of the 34 identified proteins associated with incident CVEs in CKD patients, we performed three complementary analyses. First, we evaluated pairwise Spearman correlations between the 34 proteins and 30 routinely measured blood biomarkers in the UKB, including metabolic, renal, hepatic, and inflammatory markers (**Supplementary Table 2**), to contextualize the proteins within broader physiological systems. Second, we explored potential causal relationships using summary-level two-sample Mendelian randomization (MR) results from the Proteome Phenome Atlas (https://proteome-phenome-atlas.com/) ^15,16^. We identified proteins whose levels were potentially influenced by CKD (CKD → protein) and those with evidence of causal effects on cardiovascular event risk (protein → CVE). Third, we examined tissue-specific expression profiles of the 34 proteins using RNA-seq data from the GTEx project ^17^, identifying the tissue with the highest expression (log □ [TPM + 1]) for each protein to infer potential functional relevance and tissue-level mechanisms.

### 2.4 Developing protein-based CVE risk scoring models

To evaluate the predictive utility of the 34 prioritized proteins, we developed a ProRS model by training Cox proportional hazards models with Elastic Net regularization using protein expressions. Model training was conducted in the discovery set (70%) and evaluated in the held-out evaluation set (30%), preserving the split used in the biomarker selection step. No clinical covariates were included in the ProRS to isolate the predictive contribution of protein features.

Performance was benchmarked against five conventional and genetic models (**Figure 2c**): (1) demographics and kidney function (age, sex, ethnicity, eGFR), (2) PCE variables, (3) PCE plus eGFR, (4) multi-trait polygenic risk scores (PRSs) for CHD, HF, and ischemic heart disease (IHD) with age and sex, and (5) PRSs with full PCE and eGFR. Baseline models were fit using standard Cox regression without regularization. PRSs were derived using PRS-CS-auto with FinnGen summary statistics and applied to individuals of European ancestry to ensure alignment with the discovery population ^18^.

Model performance was assessed using the C-index and time-dependent area under the curve (AUC) from 1 to 10 years. To assess risk stratification, individuals were grouped into risk categories based on model-derived scores: Very Low (<5th percentile), Low (5th–25th), Intermediate (25th–75th), High (75th–95th), and Very High (>95th), or alternatively into Low (<25th percentile), Intermediate (25th–75th), and High (>75th percentile) categories. Kaplan–Meier curves were used to visualize cumulative incidence of CVEs across these strata.

### 2.5 Statistical Analysis

All statistical analyses were conducted using Python (v3.10). Summary statistics were computed for clinical and demographic variables at baseline among UKB-PPP participants with CKD, stratified by CVE status during follow-up. Continuous variables are presented as mean ± standard deviation and were compared using Welch’s t-test. Categorical variables are presented as counts (percentages) and were compared using the Chi-squared test. Time-to-event analyses were performed using Cox proportional hazards models implemented via the lifelines package. For protein biomarker discovery, univariate Cox models were applied to each protein, adjusting for demographic and clinical covariates (age, sex, ethnicity, eGFR, SBP, DBP, HTN, BMI, DM, Smoking, and Alcohol). Proteins with nominal significance (P < 0.05) and consistent effect directions across both the discovery and evaluation sets were re-evaluated in the full cohort, and those passing a false discovery rate (FDR) threshold of q < 0.05 were retained.

ProRS were developed using Cox models with Elastic Net regularization implemented via the scikit-survival package. Hyperparameters were selected through 10-fold cross-validation within the discovery set to optimize the C-index. Conventional clinical and genetic models were fit using standard Cox models without penalization.

Model performance was evaluated using the C-index and time-dependent AUC across 1 to 10 years. Risk stratification was based on percentiles of model-derived risk scores, and Kaplan–Meier curves were used to visualize cumulative incidence across strata. All statistical tests were two-sided, and P < 0.05 was considered nominally significant unless otherwise noted.

## 3. Results

### 3.1 Baseline Characteristics

A total of 1,799 participants were selected from 44,194 individuals in the UKB-PPP based on the inclusion criteria of having CKD at baseline and no prior history of CVE. Among them, 375 CKD patients experienced one or more incident CVEs during follow-up, including CHD, ISS, HF, or cardiovascular-related mortality, while 1,424 remained CVE-free. The median follow-up time was 15.4 years (interquartile range: 14.5–16.2 years). Baseline characteristics are summarized in **Table 1**. Compared to the CVE-free group, individuals who developed CVEs were older (63.17 vs. 60.84 years), had higher SBP (143.44 vs. 140.39 mmHg), and higher BMI (29.75 vs. 28.80 kg/m^2^). Measures of kidney function and lipid profiles also differed between the groups: those with incident CVEs had lower eGFR (51.40 vs. 55.30 mL/min/1.73m^2^), TC (5.36 vs. 5.73 mmol/L), LDL (3.35 vs. 3.60 mmol/L), and HDL (1.32 vs. 1.40 mmol/L). The CVE group included a higher proportion of males (55.5% vs. 38.1%), current or former smokers (53.0% vs. 44.9%), and individuals with HTN (59.7% vs. 42.6%) or DM (13.6% vs. 6.2%). In contrast, fasting time, DBP, and alcohol consumption were similar between the two groups. The baseline characteristics highlight established cardiometabolic and renal risk factors—age, sex, blood pressure, DM, kidney function, and lipid status—as key differentiators of future CVE risk in patients with CKD.

**Table 1.** Baseline characteristics of CKD Patients with and without incident CVEs.

| Variable | CKD patients (N=1,799) | Incident CVE (N=375) | CVE free (N=1,424) | P value |
| --- | --- | --- | --- | --- |
| **Fasting Time** | 3.87 ± 2.32 | 3.90 ± 2.38 | 3.86 ± 2.31 | 8.06E-01 |
| **Age** | 61.33 ± 6.59 | 63.17 ± 5.87 | 60.84 ± 6.69 | 7.62E-11 |
| **SBP** | 141.02 ± 18.39 | 143.44 ± 18.61 | 140.39 ± 18.29 | 5.94E-03 |
| **DBP** | 82.84 ± 10.41 | 82.44 ± 11.07 | 82.95 ± 10.24 | 4.37E-01 |
| **BMI** | 29.00 ± 5.08 | 29.75 ± 5.41 | 28.80 ± 4.97 | 2.35E-03 |
| **eGFR** | 54.49 ± 13.36 | 51.40 ± 15.31 | 55.30 ± 12.69 | 8.30E-06 |
| **TC** | 5.66 ± 1.23 | 5.36 ± 1.21 | 5.73 ± 1.22 | 3.26E-07 |
| **LDL** | 3.55 ± 0.93 | 3.35 ± 0.94 | 3.60 ± 0.92 | 3.75E-06 |
| **HDL** | 1.38 ± 0.37 | 1.32 ± 0.36 | 1.40 ± 0.37 | 5.38E-04 |
| **Sex (Male)** | 751 (41.7%) | 208 (55.5%) | 543 (38.1%) | 2.01E-09 |
| **Ethnic (White)** | 1,590 (88.4%) | 337 (89.9%) | 1,253 (88.0%) | 3.59E-01 |
| **Smoking** | 841 (46.7%) | 197 (53.0%) | 637 (44.9%) | 6.35E-03 |
| **Alcohol** | 1,677 (93.2%) | 350 (93.3%) | 1,327 (93.3%) | 1.00E+00 |
| **HTN** | 830 (46.1%) | 224 (59.7%) | 606 (42.6%) | 4.14E-09 |
| **DM** | 139 (7.7%) | 51 (13.6%) | 88 (6.2%) | 2.88E-06 |

**Abbreviations:** CVE = cardiovascular event; CKD = chronic kidney disease; SBP = systolic blood pressure; DBP = diastolic blood pressure; BMI = body mass index; eGFR = estimated glomerular filtration rate; TC = total cholesterol; LDL = low-density lipoprotein cholesterol; HDL = high-density lipoprotein cholesterol; HTN = hypertension; DM = diabetes mellitus.

### 3.2 Proteins Associated with Incident CVE in CKD

From a multi-stage association analysis from a Cox proportional hazard model, we identified 34 plasma proteins significantly associated with incident CVEs in individuals with CKD. In the discovery set (264 CVE incident cases, 995 CVE-free controls), 530 proteins reached nominal significance (P < 0.05), of which 152 remained nominally significant in the evaluation set (111 incident CVE cases, 429 CVE-free controls). Thirty-eight proteins were significant in both subsets, and 34 showed consistent directions of effect (**Supplementary Figure 1**). These 34 proteins were carried forward for model development. In the full cohort (375 incident CVE cases, 1,424 CVE-free controls), all 34 proteins remained significant after FDR correction (q < 0.05). Complete summary statistics from the multi-stage association analysis are provided in **Supplementary Table 3**. Several proteins were strongly associated with increased CVE risk (e.g., ADM, SHISA5, EFCAB14, CD99L2, and PGF; HR > 2), while others were inversely associated (e.g., TMPRSS5, RET, DPP6, BCAN, and B4GAT1; HR<0.7) (**Figure 1b, Table 2**).

**Table 2.** 34 Plasma Proteins Associated With Incident Cardiovascular Events in Chronic Kidney Disease.

| Protein | UniProt | Protein Name | HR (95% CI) | P value |
| --- | --- | --- | --- | --- |
| ADM | P35318 | ADM | 2.615 [1.72,3.976] | 6.91E-06 |
| SHISA5 | Q8N114 | Protein shisa-5 | 2.581 [1.874,3.553] | 6.24E-09 |
| EFCAB14 | O75071 | EF-hand calcium-binding domain-containing protein 14 | 2.513 [1.755,3.597] | 4.82E-07 |
| CD99L2 | Q8TCZ2 | CD99 antigen-like protein 2 | 2.264 [1.516,3.38] | 6.49E-05 |
| PGF | P49763 | Placenta growth factor | 2.148 [1.579,2.921] | 1.11E-06 |
| EFEMP1 | Q12805 | EGF-containing fibulin-like extracellular matrix protein 1 | 1.994 [1.492,2.665] | 3.07E-06 |
| EDA2R | Q9HAV5 | Tumor necrosis factor receptor superfamily member 27 | 1.951 [1.53,2.488] | 7.29E-08 |
| IGLC2 | P0DOY2 | Immunoglobulin lambda constant 2 | 1.913 [1.452,2.521] | 4.03E-06 |
| CTHRC1 | Q96CG8 | Collagen triple helix repeat-containing protein 1 | 1.822 [1.324,2.507] | 2.32E-04 |
| PSAP | P07602 | Prosaposin | 1.814 [1.336,2.464] | 1.37E-04 |
| IGFBP4 | P22692 | Insulin-like growth factor-binding protein 4 | 1.805 [1.453,2.242] | 9.57E-08 |
| CLEC14A | Q86T13 | C-type lectin domain family 14 member A | 1.767 [1.316,2.372] | 1.53E-04 |
| NXPH3 | O95157 | Neurexophilin-3 | 1.685 [1.227,2.314] | 1.26E-03 |
| ANGPTL3 | Q9Y5C1 | Angiopoietin-related protein 3 | 1.618 [1.232,2.125] | 5.44E-04 |
| TNC | P24821 | Tenascin | 1.56 [1.297,1.878] | 2.46E-06 |
| EFHD1 | Q9BUP0 | EF-hand domain-containing protein D1 | 1.45 [1.186,1.772] | 2.93E-04 |
| CDSN | Q15517 | Corneodesmosin | 1.438 [1.165,1.775] | 7.07E-04 |
| CD7 | P09564 | T-cell antigen CD7 | 1.426 [1.146,1.773] | 1.44E-03 |
| MMP3 | P08254 | Stromelysin-1 | 1.409 [1.197,1.659] | 3.85E-05 |
| ACTA2 | P62736 | Actin, aortic smooth muscle | 1.375 [1.115,1.697] | 2.95E-03 |
| REG1A | P05451 | Lithostathine-1-alpha | 1.338 [1.134,1.579] | 5.68E-04 |
| MMP12 | P39900 | Macrophage metalloelastase | 1.33 [1.14,1.552] | 2.93E-04 |
| TFF2 | Q03403 | Trefoil factor 2 | 1.276 [1.102,1.477] | 1.09E-03 |
| TNFRSF10B | O14763 | Tumor necrosis factor receptor superfamily member 10B | 1.27 [1.116,1.445] | 2.84E-04 |
| NTPROBNP | NTproBNP | NT-proBNP | 1.269 [1.178,1.368] | 4.58E-10 |
| CHGA | P10645 | Chromogranin-A | 1.248 [1.147,1.357] | 2.52E-07 |
| MSLN | Q13421 | Mesothelin | 1.238 [1.079,1.421] | 2.31E-03 |
| NPPB | P16860 | Natriuretic peptides B | 1.166 [1.091,1.247] | 5.74E-06 |
| PSG1 | P11464 | Pregnancy-specific beta-1-glycoprotein 1 | 1.151 [1.055,1.256] | 1.62E-03 |
| TMPRSS5 | Q9H3S3 | Transmembrane protease serine 5 | 0.688 [0.546,0.867] | 1.54E-03 |
| RET | P07949 | Proto-oncogene tyrosine-protein kinase receptor Ret | 0.66 [0.523,0.834] | 5.00E-04 |
| DPP6 | P42658 | Dipeptidyl aminopeptidase-like protein 6 | 0.644 [0.498,0.833] | 8.03E-04 |
| BCAN | Q96GW7 | Brevican core protein | 0.617 [0.479,0.794] | 1.74E-04 |
| B4GAT1 | O43505 | Beta-1,4-glucuronyltransferase 1 | 0.517 [0.348,0.769] | 1.13E-03 |

### 3.3 Biological Characterization of Associated Proteins

The 34 proteins associated with incident CVEs in CKD patients demonstrated strong biological coherence with known cardiometabolic and renal pathways. Pearson correlation analysis with 30 standard blood biomarkers revealed significant associations (P < 0.001), particularly with cystatin C, C-reactive protein (CRP), urate, triglycerides, HDL cholesterol, and creatinine (**Figure 2a**; **Supplementary Table 4**). For example, cystatin C showed strong positive correlations with IGFBP4 (r = 0.71), CD99L2 (r = 0.43), and PGF (r = 0.63), supporting shared pathophysiologic links to kidney dysfunction and vascular injury ^19^.

To systematically evaluate the potential causal role of incident CVE-associated proteins in prevalent CKD condition, we queried the Proteome Phenome Atlas for two-sample MR evidence linking protein levels to cardiovascular outcomes and CKD (**Figure 2b; Supplementary Table 5**). Among the 34 proteins identified in our association analysis, 25 showed nominal MR evidence for a causal effect of CKD on protein levels (CKD → protein), suggesting that kidney dysfunction may influence the expression or regulation of these proteins. Of these, 11 proteins also exhibited nominal MR associations with at least one cardiovascular outcome (protein → CVE), supporting their potential role as mediators in the CKD-to-CVD pathway. For example, ANGPTL3 (angiopoietin-like 3) demonstrated consistent bidirectional evidence. Genetically predicted CKD was positively associated with ANGPTL3 levels (OR = 1.09 [95% CI: 1.02–1.17], P = 0.012). In contrast, higher genetically predicted ANGPTL3 levels were associated with reduced risk of several cardiovascular outcomes, including coronary atherosclerosis (OR = 0.51 [0.42–0.64], P = 7.2 × 10□^1^□), IHD (OR = 0.59 [0.49–0.70], P = 1.5 × 10□ □), and coronary heart disease (OR = 0.52 [0.45–0.59], P = 3.0 × 10□^21^), based on trans-protein quantitative trait locus instruments. Notably, four additional proteins (NT-proBNP, NPPB, MMP12, and EDA2R) were not significantly associated with CKD but demonstrated significant MR associations with cardiovascular outcomes, suggesting that their contribution to CVE risk may be independent of kidney function.

Tissue-specific expression analysis using GTEx RNA-seq data revealed enrichment of the 34 proteins in cardiovascular, vascular, hepatic, renal, immune, and brain tissues (**Figure 2c**; **Supplementary Table 6)**. For instance, NPPB and ACTA2 were most highly expressed in cardiac tissues; EFEMP1 and TNC in arterial tissues; and ANGPTL3 in the liver. Immune-associated proteins such as SHISA5 and IGLC2 showed peak expression in blood or lymphoid tissues.

### 3.4 Predictive Value of ProRS in UKB-PPP

To evaluate the prognostic utility of the 34 CVE-associated proteins, we developed ProRS with a Cox proportional hazards model with Elastic Net regularization using their normalized expression levels. Model performance was benchmarked against five conventional and genetic models using time-dependent discrimination, stratified risk estimation, and joint modeling. Across all models, the ProRS model achieved the highest overall C-index (0.729), outperforming PCE + eGFR (0.669), PCE alone (0.661), the combined PRS + PCE + eGFR model (0.654), and the PRS-only model (0.624) (**Supplementary Table 7**). While positive predictive values (PPVs) across all models were modest due to the low event rate, the ProRS consistently showed higher PPVs over time (e.g., PPV = 0.296 at 10 years) relative to baseline models (e.g., PCE + eGFR: PPV = 0.278; PRS-only: PPV = 0.212). Notably, the ProRS maintained high negative predictive values (NPVs ≥ 0.905) across all time points, indicating a strong ability to rule out future CVEs in low-risk individuals.

#### Discrimination across time horizons

The ProRS demonstrated superior time-dependent discrimination throughout the follow-up period, consistently outperforming all baseline models after the second year. At 10 years, the ProRS achieved an AUC of 0.725, compared to 0.692 for the best-performing clinical model (PCE + eGFR) and 0.688 for PCE alone (**Figure 3a**). The ProRS showed particularly strong performance at 3 years (AUC = 0.738) and maintained high discrimination through 9 years (AUC = 0.743). Over the full follow-up period, the ProRS achieved the highest overall AUC (0.760), notably higher than the PCE + eGFR model (0.688) and PRS-based models (≤ 0.674). These findings highlight the model’s capacity to capture long-term CVE risk that is not fully represented by traditional clinical or genetic predictors.

**Figure 3.**
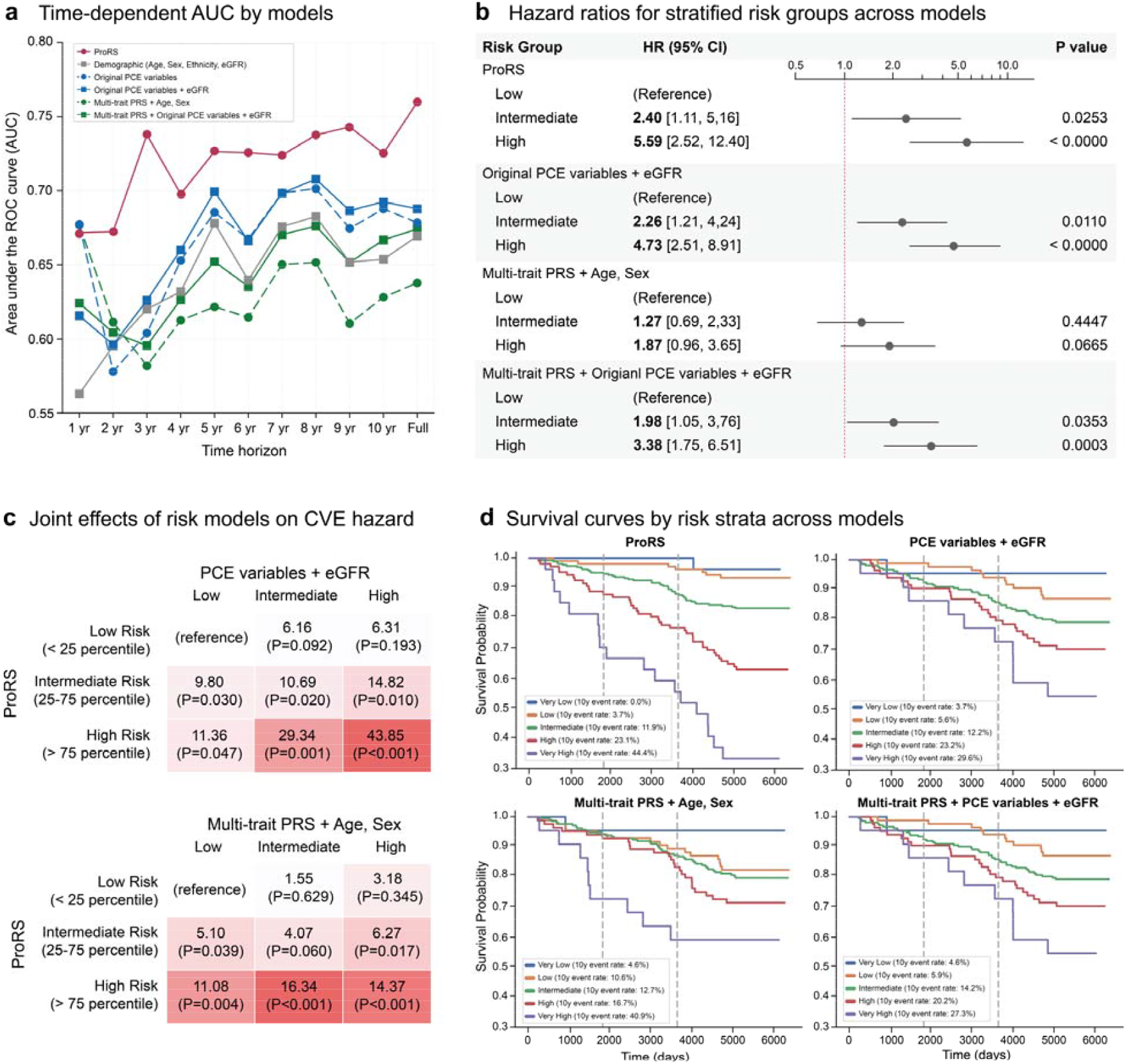
Comparative performance of the ProRS versus clinical and genetic models for cardiovascular event risk in CKD. (a) Time-dependent AUCs over a 10-year follow-up and full duration. The ProRS outperformed all clinical and genetic benchmarks, especially beyond year 3. (b) Hazard ratios across risk groups defined by model-derived percentiles (<25th, 25th– 75th, >75th), showing greater risk separation for the ProRS. (c) Joint risk stratification with clinical (PCE + eGFR) and genetic (PRS + age, sex) models. High ProRS conferred elevated hazard even when comparator models indicated low risk. (d) Kaplan– Meier curves across five percentile-based risk strata. The ProRS showed the clearest gradient, with 10-year CVE incidence ranging from 0% (very low-risk) to 44.4% (very high-risk).

#### Risk group stratification

Using model-derived scores, individuals were categorized into low (<25th percentile), intermediate (25th–75th), and high (>75th percentile) risk groups. The ProRS showed markedly improved risk separation (**Figure 3b**). Individuals in the high-risk group had a 5.59-fold increased hazard of CVE (95% CI: 2.52– 12.40, P < 0.0001) relative to the low-risk group, while the highest HRs for PCE + eGFR and PRS-based models were 3.38 or lower. Even the intermediate-risk group in the ProRS showed significantly elevated risk (HR = 2.40, 95% CI: 1.11–5.16, P = 0.025).

#### Joint stratification reveals orthogonal predictive value

Joint Cox models combining ProRS with clinical or genetic models revealed additive and independent predictive contributions (**Figure 3c**). Individuals classified as high risk by both the ProRS and PCE + eGFR models had a 43.85-fold increased hazard of CVE (P < 0.001) compared to the dual low-risk group. Among those labeled low risk by the clinical model, individuals reclassified as high risk by the ProRS still exhibited significantly increased risk (HR = 11.36, P = 0.047), whereas the converse was not observed. Similar patterns were found when combining the ProRS with multi-trait PRSs: the highest-risk group had a 14.37-fold increased hazard (P < 0.001), and those with high ProRS but low PRS still showed elevated risk (HR = 11.08, P = 0.004). These findings indicate that protein-based scores provide orthogonal and dominant prognostic information beyond existing clinical or genetic tools.

#### Survival analysis across percentiles

Further stratification into five risk groups—very low (<5th percentile), low (5th–25th), intermediate (25th–75th), high (75th–95th), and very high (>95th percentile)—demonstrated the ProRS’s ability to capture extreme risk gradients (**Figure 3d**). At 10 years, the very high-risk group (top 5%) had a 44.4% incidence of CVE, compared to 0% in the very low-risk group. In contrast, the PCE + eGFR model showed 29.6% and 3.7% event rates in the corresponding groups, while PRS-based models had weaker stratification at the lower tail (e.g., 4.6% in the bottom 5%). These results highlight the unique potential of protein-based modeling to both identify individuals at high risk and confidently exclude those at minimal risk, offering a refined tool for personalized CVE prevention in patients with CKD.

## 4. Discussion

In this study, we demonstrated that a panel of 34 plasma proteins significantly improves the prediction of incident CVEs in individuals with CKD, beyond traditional clinical and genetic risk factors. Notably, our ProRS outperformed established tools such as the Pooled Cohort Equations (PCE) and PRS, achieving higher discrimination, better precision, and more refined risk stratification across a 10-year horizon. While the CVD-PRS demonstrated strong predictive ability for individuals in the top and bottom 5% of the distribution, it failed to effectively discriminate disease risk among the remaining 90% of patients. In contrast, the clinical risk model combining PCE and eGFR was limited in its ability to identify individuals at either very high or very low risk. Interestingly, even among patients with low clinical or genetic risk factors, those with a high ProRS had a higher incidence of CVEs. Conversely, when ProRS was low, the increase in CVE incidence was minimal, even in those with high clinical or genetic risk scores. In addition, ProRS demonstrated significantly superior predictive power during the first three years, indicating its potential for early detection of CVD compared with other risk models. These findings strongly support the fact that risk stratification based on ProRS offers substantial advantages over previously established models.

Despite the high prevalence of CVEs, there have been few biomarkers for early detection of CVEs in patients with CKD. Remarkably, no specific cholesterol management guideline has been established for patients with CKD. Previous studies have reported that the sensitivity and specificity of cardiac-specific biomarkers for predicting CVD in this population are substantially reduced ^20,21^. Therefore, the development of reliable biomarkers for CVE risk prediction in patients with CKD represents an urgent and essential unmet medical need. In our results, the 10-year risk of CVEs increased progressively across the ProRS categories-very low, low, intermediate, high, and very high-demonstrating a proportional relationship with the score. This suggests that ProRS not only effectively identifies individuals at high risk of disease but also helps to reliably exclude those at very low risk. While the human genome remains unchanged from birth, proteins are dynamically regulated in response to internal and external stimuli, allowing them to more closely reflect the pathophysiological status of diseases ^22,23^. That is the reason why the proteome has been utilized not only as a biomarker for disease prediction but also as a valuable target for therapeutic intervention.

With the recent development of various cardiovascular preventive drugs, there is growing interest in evaluating their applicability to patients with CKD. Sodium-glucose cotransport 2 (SGLT2) inhibitors have been shown to reduce the incidence of CVD in CKD patients, independent of their glucose-lowering effects ^24,25^. Additionally, glucagon-like peptide-1 receptor agonists and finerenone have demonstrated cardiovascular benefits in patients with DM and CKD ^26-28^. For the drugs mentioned above, it has been reported that earlier use in the course of the disease can reduce adverse kidney events or CVEs ^28-30^. If cardiovascular-protective agents could be proactively administered to CKD patients with high ProRS, it could represent a groundbreaking opportunity to reduce CVD and mortality in this high-risk population. Furthermore, it may promote earlier and more proactive implementation of lifestyle modifications with proven cardiovascular benefits at the individual patient level ^31^.

A recent study reported a 32-protein model for predicting CVD in patients with CKD, based on proteomic data from the CRIC and ARIC cohorts ^10^. In that study as well, the ProRS outperformed the traditional PCE and eGFR model in predicting CVD outcomes. Both the CRIC and ARIC cohorts included patients with CKD or atherosclerotic diseases. Compared to the CKD population in the UKB-PPP, these cohorts were characterized by higher prevalences of DM and HTN, as well as slightly lower eGFR levels. However, the UKB-PPP CKD patients were older and had higher blood pressure on average. In addition, the number of proteins and the proteomic platforms were different between the two studies. Perhaps as a result, apart from NTproBNP and EGF-containing fibulin-like extracellular matrix protein 1 (EFEMP1 or fibulin), the specific proteins with significant associations with CVD were different.

In our study, 25 out of the 34 proteins were found to be altered in the presence of CKD, as determined by Mendelian randomization analysis. In the clustered heatmap, many of these proteins were also associated with blood biomarkers related to kidney function. Of the 24 proteins, 10 are thought to play causal roles in at least one of the three diseases: IHD, HF, and CAD. Chronic sterile inflammation is a shared pathophysiological mechanism between CKD and CVD. Anti-inflammatory therapies targeting interleukin-1 (IL-1) or interleukin-6 (IL-6) have been shown in some studies to reduce CVEs ^32-34^. Among the 10 proteins, 5 proteins (TNFRSF10B, MMP3, TFF2, ANGPTL3, MMP12) were associated with inflammation and immune regulation ^35-39^. Intracellular ANGPT3 showed to inhibit IL-1β-induced NF-κB activation by disrupting the assembly of the IL-1 receptor complex ^40^. Conversely, treatment with ANGPTL3 in macrophage cells increases the expression of IL-1β and tumor necrosis factor-α, suggesting that ANGPTL3 may play a regulatory role in the IL-1 pathways ^41^. In TFF2-deficient macrophages, the expressions of IL-6 and IL-1β were reduced following lipopolysaccharide stimulation. This finding suggested that TFF2 may modulate the functional interaction between IL-1 and IL-6 signaling pathways ^42^. Inflammation in CKD is closely linked to cardiac fibrosis and myocardial remodeling. Three proteins (ACTA2, MMP3, MMP12) were associated with fibrosis. ACTA2, which encodes alpha-smooth muscle actin, is considered as a marker of myofibroblasts, key player in the development of cardiac fibrosis ^43,44^. EFEMP1 is associated with vascular remodeling in the hypertensive rat model and recognized as a marker for aging in dementia ^45,46^. Considering the biological functions of the identified proteins, the most prominent signals were related to inflammation, immune regulation, and aging, which are key pathophysiologic processes in cardiorenal syndrome ^6^. These findings suggest that the proteins may serve not only as biomarkers but also as potential therapeutic targets in the future.

Our study has several limitations. First, we were not able to perform external validation. Although large-scale proteomic datasets have recently become available, their use is still limited. Nevertheless, our study produced robust findings through internal validation and meta-analysis within the UKB-PPP. Second, we were not able to include albuminuria data due to its limited availability in the UKB-PPP. Albuminuria is a well-established risk factor for predicting CVD in patients with CKD ^47^. However, considering that the UKB is a population-based cohort consisting largely of healthy individuals, the CKD patients included in our study were likely to be in earlier stages of CKD with a relatively low prevalence of DM, suggesting a lower burden of albuminuria. Despite the exclusion of albuminuria, the proteomic risk scores demonstrated strong predictive performance.

Despite these limitations, our results demonstrate the strong and independent predictive value of circulating plasma proteins for CVEs in CKD and provide a scalable framework for developing proteomics-driven risk assessment tools. These protein markers, grounded in disease-relevant biology, offer a path toward more precise and personalized cardiovascular care in a population for whom improved prognostic tools are urgently needed.

## Data Availability

All data produced in the present study are available upon reasonable request to the authors

## Acknowledgements

We acknowledge all the participants of the UK Biobank. The use of the UK Biobank resources was approved under Application Number 32133.

