## Supplementary Material for "Protein risk scores enable precise prediction of cardiovascular events in chronic kidney disease patients"

**Supplementary Figures**

Supplementary Figure 1. Multi-stage identification of plasma proteins associated with incident CVEs in CKD

**Supplementary Tables**

Supplementary Table 1. Phenotyping definitions for disease conditions in this study

Supplementary Table 2. Summary statistics of 30 blood biomarkers in the UK Biobank cohort.

Supplementary Table 3. Full summary statistics from multi-stage association analysis of protein expression and CVE risk.

Supplementary Table 4. Pairwise correlations between 30 clinical blood biomarkers and the 34 CVE-associated proteins

Supplementary Table 5. Two-sample Mendelian Randomization results linking protein levels to cardiovascular outcomes and CKD

Supplementary Table 6. Tissue-specific expression of the 34 proteins associated with incident CVEs in CKD.

Supplementary Table 7. Predictive performance of risk models for CVE in CKD

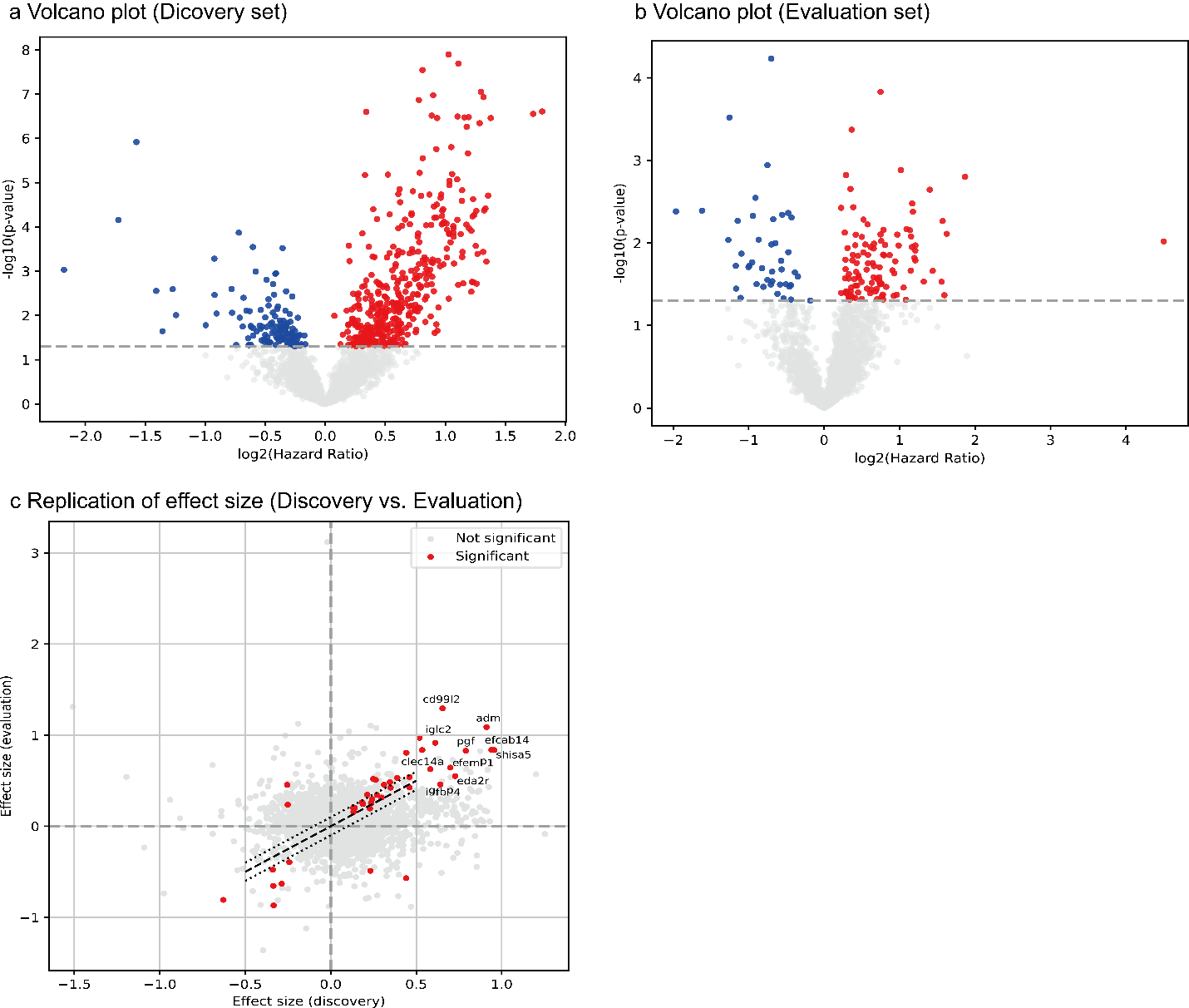

**Supplementary Figure 1 | Multi-stage identification of plasma proteins associated with incident CVEs in CKD.**
(a) Volcano plot showing protein associations with incident CVEs in the discovery set (264 cases, 995 controls). Red and blue dots indicate proteins with nominal significance (P < 0.05) and positive or negative log₂ hazard ratios, respectively. (b) Volcano plot of the evaluation set (111 cases, 429 controls), using the same significance threshold. (c) Replication of effect sizes for all proteins between discovery and evaluation sets. Each dot represents a protein, with red dots indicating proteins that were nominally significant in both subsets and had consistent effect directions. The 34 proteins highlighted in this analysis were selected for downstream model development.

**Supplementary Table 1 | Phenotyping definitions for disease conditions in this study.** This table summarizes the phenotype definitions used to ascertain disease conditions from the UK Biobank dataset. Each phenotype is mapped to its corresponding UKB Data-Field and associated codes from ICD-9, ICD-10, and OPCS classification systems.

| **Diesease phenotypes** | **UKB Data-Field** | **Code (UKB; ICD9; ICD10; OPCS code)** |
| --- | --- | --- |
| Chronic Kidney Disease | non-cancer illness, self-report (20002, 20008) | kidney nephropathy (1519); diabetic nephropathy (1607); renal/kidney failure (1192); renal failure not requiring dialysis (1194) |
| Chronic Kidney Disease | first occurrence | N18; N19 |
| Chronic Kidney Disease | diagnoses - ICD10 (41270, 41280) | E112; E132; E142; E12; E13; N18; N19; N271 |
| Chronic Kidney Disease | blood assay (30700) | CKD EPI (cystatin C) eGFR < 60 |
| End-Stage Renal Disease | Operation code, self-report (20004, 20010) | fistua for dialysis (1476); dialysis access surgery (1580); HD access/fistula surgery (1581); PD access surgery (1582) |
| End-Stage Renal Disease | non-cancer illness, self-report (20002, 20008) | renal failure requiring dialysis (1193) |
| End-Stage Renal Disease | diagnoses - ICD9 (41271, 41281) | V451; V568 |
| End-Stage Renal Disease | diagnoses - ICD10 (41270, 41280) | N180; N185; Q601; T824; Z49; Z992; Y602 |
| End-Stage Renal Disease | Operative procedures - OPCS4 | L74; X40; X41; X42; M203; M172 |
| End-Stage Renal Disease | Operative procedures - OPCS3 | 401.3; 889.1 |
| End-Stage Renal Disease | Algorithm-defined outcome | date of end stage renal disease report (42026, 42027) |
| Kidney Transplant | Operation code, self-report (20004, 20010) | renal/kidney transplant (1195) |
| Kidney Transplant | diagnoses - ICD9 (41271, 41281) | V420 |
| Kidney Transplant | diagnoses - ICD10 (41270, 41280) | N165; T861; Z94; Y841 |
| Kidney Transplant | Operative procedures - OPCS4 | M01; M084; M174; M178; M179 |
| Kidney Transplant | Operative procedures - OPCS3 | 566 |
| Cardiovascular Event | self-reported medical conditions | Age angina diagnosed (3627); Age heart attack diagnosed(3894) |
| Cardiovascular Event | non-cancer illness, self-report (20002, 20008) | 1074; 1075; 1081; 1082; 1583; 1094 |
| Cardiovascular Event | first occurrence | 131296; 131298; 131300; 131302; 131304; 131306; 131362; 131364; 131368 |
| Cardiovascular Event | diagnoses - ICD9 (41271, 41281) | 4109; 4119; 4129; 4140; 4148; 4149; 4309; 4319; 4321; 4320; 4331; 4339; 4349; 4359; 4369; 4370; 4371; 4378; 4379; 4389; 4280; 4281; 4289 |
| Cardiovascular Event | diagnoses - ICD10 (41270, 41280) | I20; I21; I22; I23; I24; I25; Z951; I60; I61; I62; I63; I64; I65; I66; I67; I68; I69; I500; I501; I509; I13; I50; I517; I518; I519; I52 |
| Cardiovascular Event | Operative procedures - OPCS4 | K40; K41; K42; K43; K44; K45; K46; K47; K48 |
| Cardiovascular Event | Operative procedures - OPCS3 | 3041; 3042; 3043 |
| Cardiovascular Event | Operation code, self-report (20004, 20010) | 1070; 1095 |
| Cardiovascular Event | Algorithm-defined outcome | Date of myocardial infarction (42000); Date of STEMI (42002); Date of NSTEMI (42004); Date of stroke (42006); Date of ischaemic stroke (42008); Date of intracerebral haemorrhage (42010); Date of subarachnoid haemorrhage (42012) |
| Cardiovascular Event | primary cause of death: ICD10 | I13; I146; I20; I21; I22; I23; I24; I25; I50; I517; I518; I519; I52; I60; I61; I62; I63; I64; I65; I66; I67; I68; I69; Z951 |
| Hypertension | non-cancer illness, self-report (20002, 20008) | 1065; 1072 |
| Hypertension | self-reported medical conditions | high blood pressurer (6150) |
| Hypertension | first occurrence | I10; I11; I12; I13; I15 |
| Hypertension | blood assay (4080) | ≥ 140 mmHg |
| Hypertension | blood assay (4079) | ≥ 90 mmHg |
| Hypertension | diagnoses - ICD10 (41270, 41280) | I10; I11; I12; I13; I14; I15 |
| Diabetes Mellitus | non-cancer illness, self-report (20002, 20008) | 1220; 1222; 1223 |
| Diabetes Mellitus | first occurrence | E10; E11; E14 |
| Diabetes Mellitus | diagnoses - ICD10 (41270, 41280) | E10; E11; E13; E14 |
| Diabetes Mellitus | diagnoses - ICD9 (41271, 41281) | 250 |
| Diabetes Mellitus | treatment/medication code (20003) | insulin (1140884066); metformin (1140884600, 1141189090); sulfonylurea (1141152590, 1140874744, 1140874718, 1141156984); acarbose (1140868902); meglitinide (1141168660, 1141173882); glitazones (1141153254, 1141171646, 1141177600); combination (1141189090, 1141189094) |
| Diabetes Mellitus | blood assay (30750) | glycated hemoglobin (HbA1c) ≥ 6.5% (48 mmol/mol or over) |

**Supplementary Table 2 | Summary statistics of 30 blood biomarkers in the UK Biobank cohort.** For each biomarker, mean and standard deviation (std) are reported across the total population, individuals with incident cardiovascular events (CVE), and those who remained CVE-free. These biomarkers were used to explore associations with proteomic risk scores and stratified CVE risk in chronic kidney disease patients

| **Blood Biomarker** | **Total** | | **CVE incident** | | **CVE free** | |
| --- | --- | --- | --- | --- | --- | --- |
|  | **mean** | **std** | **mean** | **std** | **mean** | **std** |
| ALT | 23.77 | 14.81 | 23.17 | 11.54 | 23.93 | 15.56 |
| Albumin | 44.49 | 2.86 | 43.90 | 2.80 | 44.65 | 2.85 |
| ALP | 90.09 | 27.42 | 91.41 | 28.74 | 89.74 | 27.07 |
| ApoA | 1.51 | 0.27 | 1.46 | 0.25 | 1.52 | 0.27 |
| ApoB | 1.03 | 0.25 | 0.98 | 0.25 | 1.05 | 0.25 |
| AST | 27.35 | 11.32 | 27.27 | 11.08 | 27.37 | 11.38 |
| CRP | 3.77 | 6.11 | 3.60 | 4.24 | 3.81 | 6.51 |
| Calcium | 2.39 | 0.10 | 2.37 | 0.10 | 2.40 | 0.10 |
| Cholesterol | 5.66 | 1.23 | 5.36 | 1.21 | 5.73 | 1.22 |
| Creatinine | 96.39 | 25.28 | 103.43 | 38.51 | 94.54 | 20.04 |
| Cystatin C | 1.17 | 0.27 | 1.29 | 0.37 | 1.14 | 0.23 |
| Direct bilirubin | 1.86 | 0.83 | 1.88 | 0.87 | 1.85 | 0.82 |
| GGT | 41.24 | 47.40 | 43.40 | 48.58 | 40.68 | 47.08 |
| Glucose | 5.27 | 1.48 | 5.47 | 2.18 | 5.22 | 1.23 |
| HbA1c | 37.54 | 7.39 | 38.52 | 9.19 | 37.29 | 6.83 |
| HDL | 1.38 | 0.37 | 1.32 | 0.36 | 1.40 | 0.37 |
| IGF-1 | 20.95 | 6.24 | 20.71 | 6.77 | 21.02 | 6.10 |
| LDL | 3.55 | 0.93 | 3.35 | 0.94 | 3.61 | 0.92 |
| Lipoprotein A | 47.78 | 52.35 | 45.71 | 51.75 | 48.30 | 52.51 |
| Oestradiol | 308.35 | 234.57 | 259.79 | 117.24 | 320.48 | 254.56 |
| Phosphate | 1.18 | 0.17 | 1.17 | 0.17 | 1.18 | 0.16 |
| Rheumatoid factor | 26.00 | 19.27 | 28.95 | 19.55 | 25.26 | 19.19 |
| SHBG | 49.19 | 25.10 | 48.46 | 22.48 | 49.38 | 25.74 |
| Testosterone | 6.12 | 5.93 | 7.47 | 5.96 | 5.74 | 5.86 |
| Total bilirubin | 9.06 | 4.37 | 8.93 | 4.46 | 9.10 | 4.35 |
| Total protein | 72.65 | 4.42 | 72.35 | 4.68 | 72.72 | 4.35 |
| Triglycerides | 1.95 | 1.04 | 2.03 | 1.14 | 1.92 | 1.01 |
| Urate | 358.80 | 81.63 | 374.30 | 83.65 | 354.71 | 80.63 |
| Urea | 6.75 | 2.01 | 7.36 | 2.79 | 6.59 | 1.71 |
| Vitamin D | 49.20 | 21.97 | 46.36 | 22.68 | 49.96 | 21.73 |

**Supplementary Table 3 | Full summary statistics from multi-stage association analysis of protein expression and CVE risk.** Results of Cox proportional hazards models evaluating the association between plasma protein levels and incident cardiovascular events among CKD patients. The table includes effect size estimates (coefficients, hazard ratios, and 95% confidence intervals), raw and FDR-adjusted p-values, and log-transformed metrics for each of the 2,920 proteins across the discovery set (70%), evaluation set (30%), and combined meta-analysis (100%).

| **Discovery Set (70%)** | | | | | | | | | |
| --- | --- | --- | --- | --- | --- | --- | --- | --- | --- |
| **Protein** | **UniProt** | **coef** | **HR** | **CI_lower** | **CI_upper** | **p** | **FDR** | **log2HR** | **-log10FDR** |
| **acta2** | P62736 | 0.2629 | 1.3007 | 1.0156 | 1.6658 | 0.0373 | 0.2356 | 0.3793 | 0.6278 |
| **adm** | P35318 | 0.9114 | 2.4877 | 1.5068 | 4.1071 | 0.0004 | 0.0107 | 1.3148 | 1.9710 |
| **angptl3** | Q9Y5C1 | 0.4603 | 1.5845 | 1.1465 | 2.1898 | 0.0053 | 0.0700 | 0.6640 | 1.1546 |
| **b4gat1** | O43505 | -0.6279 | 0.5337 | 0.3331 | 0.8551 | 0.0090 | 0.0966 | -0.9059 | 1.0150 |
| **bcan** | Q96GW7 | -0.3339 | 0.7161 | 0.5290 | 0.9693 | 0.0306 | 0.2119 | -0.4818 | 0.6738 |
| **cd7** | P09564 | 0.3500 | 1.4191 | 1.0931 | 1.8424 | 0.0086 | 0.0938 | 0.5050 | 1.0276 |
| **cd99l2** | Q8TCZ2 | 0.6536 | 1.9224 | 1.1995 | 3.0809 | 0.0066 | 0.0800 | 0.9429 | 1.0970 |
| **cdsn** | Q15517 | 0.3113 | 1.3653 | 1.0627 | 1.7539 | 0.0148 | 0.1398 | 0.4492 | 0.8544 |
| **chga** | P10645 | 0.2290 | 1.2574 | 1.1380 | 1.3892 | 0.0000 | 0.0007 | 0.3304 | 3.1359 |
| **clec14a** | Q86T13 | 0.5821 | 1.7898 | 1.2501 | 2.5626 | 0.0015 | 0.0288 | 0.8398 | 1.5411 |
| **cthrc1** | Q96CG8 | 0.5192 | 1.6807 | 1.1462 | 2.4645 | 0.0078 | 0.0881 | 0.7490 | 1.0548 |
| **dpp6** | P42658 | -0.3355 | 0.7150 | 0.5205 | 0.9821 | 0.0383 | 0.2390 | -0.4840 | 0.6217 |
| **eda2r** | Q9HAV5 | 0.7275 | 2.0698 | 1.5380 | 2.7855 | 0.0000 | 0.0002 | 1.0495 | 3.6377 |
| **efcab14** | O75071 | 0.9402 | 2.5606 | 1.6630 | 3.9427 | 0.0000 | 0.0015 | 1.3565 | 2.8167 |
| **efemp1** | Q12805 | 0.6996 | 2.0129 | 1.4204 | 2.8527 | 0.0001 | 0.0038 | 1.0093 | 2.4229 |
| **efhd1** | Q9BUP0 | 0.3458 | 1.4132 | 1.1184 | 1.7857 | 0.0038 | 0.0542 | 0.4989 | 1.2663 |
| **igfbp4** | P22692 | 0.6410 | 1.8983 | 1.4596 | 2.4689 | 0.0000 | 0.0002 | 0.9247 | 3.6136 |
| **iglc2** | P0DOY2 | 0.6112 | 1.8427 | 1.3570 | 2.5021 | 0.0001 | 0.0038 | 0.8818 | 2.4189 |
| **mmp12** | P39900 | 0.2971 | 1.3459 | 1.1175 | 1.6211 | 0.0017 | 0.0327 | 0.4286 | 1.4855 |
| **mmp3** | P08254 | 0.2486 | 1.2822 | 1.0419 | 1.5780 | 0.0189 | 0.1568 | 0.3587 | 0.8047 |
| **msln** | Q13421 | 0.1832 | 1.2011 | 1.0169 | 1.4187 | 0.0310 | 0.2127 | 0.2644 | 0.6723 |
| **nppb** | P16860 | 0.1398 | 1.1500 | 1.0619 | 1.2455 | 0.0006 | 0.0145 | 0.2017 | 1.8381 |
| **ntprobnp** | NTproBNP | 0.2361 | 1.2663 | 1.1576 | 1.3852 | 0.0000 | 0.0001 | 0.3406 | 4.1978 |
| **nxph3** | O95157 | 0.4422 | 1.5561 | 1.0592 | 2.2860 | 0.0242 | 0.1831 | 0.6379 | 0.7373 |
| **pgf** | P49763 | 0.7901 | 2.2036 | 1.5412 | 3.1507 | 0.0000 | 0.0014 | 1.1399 | 2.8686 |
| **psap** | P07602 | 0.5350 | 1.7075 | 1.1933 | 2.4433 | 0.0034 | 0.0513 | 0.7719 | 1.2903 |
| **psg1** | P11464 | 0.1333 | 1.1426 | 1.0246 | 1.2740 | 0.0165 | 0.1486 | 0.1923 | 0.8279 |
| **reg1a** | P05451 | 0.2706 | 1.3107 | 1.0818 | 1.5880 | 0.0057 | 0.0736 | 0.3903 | 1.1329 |
| **ret** | P07949 | -0.2872 | 0.7504 | 0.5657 | 0.9954 | 0.0464 | 0.2612 | -0.4143 | 0.5830 |
| **shisa5** | Q8N114 | 0.9555 | 2.6001 | 1.8004 | 3.7549 | 0.0000 | 0.0001 | 1.3785 | 4.1978 |
| **tff2** | Q03403 | 0.2133 | 1.2378 | 1.0331 | 1.4830 | 0.0207 | 0.1653 | 0.3078 | 0.7818 |
| **tmprss5** | Q9H3S3 | -0.3391 | 0.7124 | 0.5378 | 0.9438 | 0.0181 | 0.1544 | -0.4892 | 0.8113 |
| **tnc** | P24821 | 0.4603 | 1.5846 | 1.2633 | 1.9875 | 0.0001 | 0.0033 | 0.6641 | 2.4837 |
| **tnfrsf10b** | O14763 | 0.2420 | 1.2738 | 1.0860 | 1.4941 | 0.0029 | 0.0458 | 0.3492 | 1.3387 |
| **Evaluation Set (30%)** | | | | | | | | | |
| **Protein** | **UniProt** | **coef** | **HR** | **CI_lower** | **CI_upper** | **p** | **FDR** | **log2HR** | **-log10FDR** |
| **acta2** | P62736 | 0.50862 | 1.66299 | 1.08469 | 2.54961 | 0.01966 | 0.74552 | 0.73378 | 0.12754 |
| **adm** | P35318 | 1.08818 | 2.96886 | 1.37868 | 6.39314 | 0.00543 | 0.63392 | 1.56991 | 0.19797 |
| **angptl3** | Q9Y5C1 | 0.53949 | 1.71513 | 1.01916 | 2.88634 | 0.04221 | 0.82803 | 0.77831 | 0.08195 |
| **b4gat1** | O43505 | -0.80835 | 0.44559 | 0.20952 | 0.94764 | 0.03576 | 0.82803 | -1.16621 | 0.08195 |
| **bcan** | Q96GW7 | -0.86954 | 0.41914 | 0.26151 | 0.67179 | 0.00030 | 0.29487 | -1.25449 | 0.53037 |
| **cd7** | P09564 | 0.42319 | 1.52683 | 1.01838 | 2.28912 | 0.04055 | 0.82803 | 0.61054 | 0.08195 |
| **cd99l2** | Q8TCZ2 | 1.29405 | 3.64754 | 1.63429 | 8.14083 | 0.00158 | 0.57754 | 1.86692 | 0.23842 |
| **cdsn** | Q15517 | 0.45542 | 1.57684 | 1.05587 | 2.35487 | 0.02604 | 0.81335 | 0.65704 | 0.08972 |
| **chga** | P10645 | 0.19524 | 1.21560 | 1.03711 | 1.42482 | 0.01597 | 0.71537 | 0.28167 | 0.14547 |
| **clec14a** | Q86T13 | 0.62725 | 1.87245 | 1.09699 | 3.19606 | 0.02149 | 0.75599 | 0.90492 | 0.12148 |
| **cthrc1** | Q96CG8 | 0.97023 | 2.63856 | 1.41527 | 4.91920 | 0.00227 | 0.63392 | 1.39975 | 0.19797 |
| **dpp6** | P42658 | -0.65436 | 0.51977 | 0.33017 | 0.81825 | 0.00471 | 0.63392 | -0.94405 | 0.19797 |
| **eda2r** | Q9HAV5 | 0.55042 | 1.73398 | 1.11722 | 2.69123 | 0.01412 | 0.67576 | 0.79409 | 0.17021 |
| **efcab14** | O75071 | 0.83779 | 2.31125 | 1.21449 | 4.39845 | 0.01071 | 0.65386 | 1.20868 | 0.18451 |
| **efemp1** | Q12805 | 0.64394 | 1.90397 | 1.09112 | 3.32238 | 0.02339 | 0.77622 | 0.92901 | 0.11001 |
| **efhd1** | Q9BUP0 | 0.48274 | 1.62051 | 1.03222 | 2.54408 | 0.03593 | 0.82803 | 0.69645 | 0.08195 |
| **igfbp4** | P22692 | 0.45807 | 1.58102 | 1.07785 | 2.31908 | 0.01910 | 0.74374 | 0.66086 | 0.12858 |
| **iglc2** | P0DOY2 | 0.91518 | 2.49722 | 1.09588 | 5.69047 | 0.02942 | 0.82803 | 1.32032 | 0.08195 |
| **mmp12** | P39900 | 0.31511 | 1.37042 | 1.04096 | 1.80414 | 0.02470 | 0.80129 | 0.45461 | 0.09621 |
| **mmp3** | P08254 | 0.51859 | 1.67965 | 1.28492 | 2.19565 | 0.00015 | 0.21634 | 0.74816 | 0.66487 |
| **msln** | Q13421 | 0.26299 | 1.30081 | 1.00547 | 1.68290 | 0.04534 | 0.82803 | 0.37941 | 0.08195 |
| **nppb** | P16860 | 0.20121 | 1.22288 | 1.07997 | 1.38470 | 0.00151 | 0.57754 | 0.29029 | 0.23842 |
| **ntprobnp** | NTproBNP | 0.25316 | 1.28809 | 1.11890 | 1.48287 | 0.00043 | 0.31068 | 0.36524 | 0.50768 |
| **nxph3** | O95157 | 0.80656 | 2.24019 | 1.20103 | 4.17847 | 0.01122 | 0.65504 | 1.16362 | 0.18373 |
| **pgf** | P49763 | 0.82890 | 2.29081 | 1.17322 | 4.47298 | 0.01519 | 0.70392 | 1.19586 | 0.15248 |
| **psap** | P07602 | 0.83713 | 2.30973 | 1.16673 | 4.57247 | 0.01628 | 0.71537 | 1.20772 | 0.14547 |
| **psg1** | P11464 | 0.15633 | 1.16921 | 1.00686 | 1.35775 | 0.04041 | 0.82803 | 0.22554 | 0.08195 |
| **reg1a** | P05451 | 0.34396 | 1.41052 | 1.00358 | 1.98247 | 0.04765 | 0.82803 | 0.49622 | 0.08195 |
| **ret** | P07949 | -0.63082 | 0.53216 | 0.35164 | 0.80535 | 0.00285 | 0.63392 | -0.91008 | 0.19797 |
| **shisa5** | Q8N114 | 0.83651 | 2.30831 | 1.19856 | 4.44557 | 0.01236 | 0.67576 | 1.20683 | 0.17021 |
| **tff2** | Q03403 | 0.34650 | 1.41411 | 1.09263 | 1.83018 | 0.00846 | 0.65386 | 0.49989 | 0.18451 |
| **tmprss5** | Q9H3S3 | -0.47752 | 0.62032 | 0.41190 | 0.93421 | 0.02227 | 0.76081 | -0.68891 | 0.11872 |
| **tnc** | P24821 | 0.42718 | 1.53292 | 1.10191 | 2.13253 | 0.01121 | 0.65504 | 0.61629 | 0.18373 |
| **tnfrsf10b** | O14763 | 0.29859 | 1.34796 | 1.05368 | 1.72443 | 0.01750 | 0.71537 | 0.43078 | 0.14547 |
| **Meta-Analysis (100%)** | | | | | | | | | |
| **Protein** | **UniProt** | **coef** | **HR** | **CI_lower** | **CI_upper** | **p** | **FDR** | **log2HR** | **-log10FDR** |
| **acta2** | P62736 | 0.3187 | 1.3753 | 1.1147 | 1.6969 | 0.0030 | 0.0490 | 0.4597 | 1.3098 |
| **adm** | P35318 | 0.9613 | 2.6152 | 1.7199 | 3.9764 | 0.0000 | 0.0007 | 1.3869 | 3.1575 |
| **angptl3** | Q9Y5C1 | 0.4810 | 1.6177 | 1.2317 | 2.1247 | 0.0005 | 0.0146 | 0.6940 | 1.8365 |
| **b4gat1** | O43505 | -0.6590 | 0.5174 | 0.3480 | 0.7692 | 0.0011 | 0.0247 | -0.9507 | 1.6067 |
| **bcan** | Q96GW7 | -0.4830 | 0.6170 | 0.4794 | 0.7939 | 0.0002 | 0.0069 | -0.6968 | 2.1623 |
| **cd7** | P09564 | 0.3546 | 1.4256 | 1.1463 | 1.7729 | 0.0014 | 0.0295 | 0.5116 | 1.5297 |
| **cd99l2** | Q8TCZ2 | 0.8170 | 2.2637 | 1.5160 | 3.3802 | 0.0001 | 0.0036 | 1.1787 | 2.4463 |
| **cdsn** | Q15517 | 0.3633 | 1.4381 | 1.1654 | 1.7746 | 0.0007 | 0.0179 | 0.5242 | 1.7480 |
| **chga** | P10645 | 0.2214 | 1.2478 | 1.1471 | 1.3574 | 0.0000 | 0.0001 | 0.3194 | 4.1359 |
| **clec14a** | Q86T13 | 0.5692 | 1.7669 | 1.3160 | 2.3723 | 0.0002 | 0.0063 | 0.8212 | 2.2015 |
| **cthrc1** | Q96CG8 | 0.5997 | 1.8216 | 1.3237 | 2.5067 | 0.0002 | 0.0084 | 0.8652 | 2.0783 |
| **dpp6** | P42658 | -0.4402 | 0.6439 | 0.4978 | 0.8329 | 0.0008 | 0.0194 | -0.6351 | 1.7128 |
| **eda2r** | Q9HAV5 | 0.6683 | 1.9509 | 1.5296 | 2.4883 | 0.0000 | 0.0000 | 0.9642 | 4.3715 |
| **efcab14** | O75071 | 0.9213 | 2.5126 | 1.7552 | 3.5969 | 0.0000 | 0.0001 | 1.3292 | 3.9658 |
| **efemp1** | Q12805 | 0.6902 | 1.9942 | 1.4923 | 2.6649 | 0.0000 | 0.0004 | 0.9958 | 3.3699 |
| **efhd1** | Q9BUP0 | 0.3713 | 1.4497 | 1.1857 | 1.7723 | 0.0003 | 0.0095 | 0.5357 | 2.0213 |
| **igfbp4** | P22692 | 0.5904 | 1.8047 | 1.4528 | 2.2418 | 0.0000 | 0.0000 | 0.8517 | 4.3715 |
| **iglc2** | P0DOY2 | 0.6489 | 1.9134 | 1.4521 | 2.5214 | 0.0000 | 0.0005 | 0.9362 | 3.2908 |
| **mmp12** | P39900 | 0.2850 | 1.3297 | 1.1397 | 1.5515 | 0.0003 | 0.0095 | 0.4111 | 2.0213 |
| **mmp3** | P08254 | 0.3429 | 1.4090 | 1.1968 | 1.6589 | 0.0000 | 0.0026 | 0.4947 | 2.5825 |
| **msln** | Q13421 | 0.2138 | 1.2384 | 1.0793 | 1.4209 | 0.0023 | 0.0403 | 0.3085 | 1.3944 |
| **nppb** | P16860 | 0.1540 | 1.1665 | 1.0914 | 1.2467 | 0.0000 | 0.0007 | 0.2221 | 3.1826 |
| **ntprobnp** | NTproBNP | 0.2384 | 1.2692 | 1.1776 | 1.3680 | 0.0000 | 0.0000 | 0.3440 | 5.8740 |
| **nxph3** | O95157 | 0.5217 | 1.6849 | 1.2270 | 2.3137 | 0.0013 | 0.0265 | 0.7527 | 1.5760 |
| **pgf** | P49763 | 0.7644 | 2.1476 | 1.5789 | 2.9211 | 0.0000 | 0.0002 | 1.1027 | 3.6634 |
| **psap** | P07602 | 0.5956 | 1.8141 | 1.3358 | 2.4636 | 0.0001 | 0.0059 | 0.8592 | 2.2318 |
| **psg1** | P11464 | 0.1404 | 1.1508 | 1.0546 | 1.2558 | 0.0016 | 0.0319 | 0.2026 | 1.4961 |
| **reg1a** | P05451 | 0.2913 | 1.3382 | 1.1339 | 1.5793 | 0.0006 | 0.0149 | 0.4203 | 1.8263 |
| **ret** | P07949 | -0.4149 | 0.6604 | 0.5229 | 0.8342 | 0.0005 | 0.0135 | -0.5985 | 1.8688 |
| **shisa5** | Q8N114 | 0.9480 | 2.5807 | 1.8743 | 3.5532 | 0.0000 | 0.0000 | 1.3677 | 5.0402 |
| **tff2** | Q03403 | 0.2438 | 1.2760 | 1.1024 | 1.4770 | 0.0011 | 0.0242 | 0.3517 | 1.6154 |
| **tmprss5** | Q9H3S3 | -0.3735 | 0.6883 | 0.5463 | 0.8673 | 0.0015 | 0.0308 | -0.5389 | 1.5115 |
| **tnc** | P24821 | 0.4449 | 1.5604 | 1.2967 | 1.8776 | 0.0000 | 0.0004 | 0.6419 | 3.4453 |
| **tnfrsf10b** | O14763 | 0.2390 | 1.2700 | 1.1162 | 1.4450 | 0.0003 | 0.0095 | 0.3448 | 2.0213 |

**Supplementary Table 4 | Pairwise correlations between 30 clinical blood biomarkers and the 34 CVE-associated proteins.** Pearson correlation coefficients were computed using normalized values of blood biomarkers and protein expression levels in CKD patients at baseline. This analysis evaluates the degree of association between conventional clinical markers and proteomic features selected in the final model.

|  | **Protein** | **Blood biomarker** | **coefficient** |  | **Protein** | **Blood biomarker** | **coefficient** |  | **Protein** | **Blood biomarker** | **coefficient** |
| --- | --- | --- | --- | --- | --- | --- | --- | --- | --- | --- | --- |
| 1 | acta2 | ALT | -0.082 | 211 | cthrc1 | Vitamin D | -0.057 | 421 | ntprobnp | HDL | 0.102 |
| 2 | acta2 | Albumin | -0.124 | 212 | dpp6 | Albumin | 0.146 | 422 | ntprobnp | IGF-1 | -0.162 |
| 3 | acta2 | ALP | 0.09 | 213 | dpp6 | ALP | 0.059 | 423 | ntprobnp | LDL | -0.092 |
| 4 | acta2 | ApoA | 0.054 | 214 | dpp6 | ApoA | 0.151 | 424 | ntprobnp | Phosphate | 0.079 |
| 5 | acta2 | CRP | 0.128 | 215 | dpp6 | AST | 0.256 | 425 | ntprobnp | SHBG | 0.278 |
| 6 | acta2 | Creatinine | 0.075 | 216 | dpp6 | CRP | -0.169 | 426 | ntprobnp | Testosterone | -0.106 |
| 7 | acta2 | Cystatin C | 0.444 | 217 | dpp6 | Calcium | 0.127 | 427 | ntprobnp | Total protein | -0.094 |
| 8 | acta2 | Glucose | 0.061 | 218 | dpp6 | Cholesterol | 0.048 | 428 | ntprobnp | Triglycerides | -0.135 |
| 9 | acta2 | HbA1c | 0.11 | 219 | dpp6 | Creatinine | -0.127 | 429 | ntprobnp | Urea | 0.135 |
| 10 | acta2 | IGF-1 | -0.107 | 220 | dpp6 | Cystatin C | -0.115 | 430 | ntprobnp | Vitamin D | 0.088 |
| 11 | acta2 | SHBG | 0.083 | 221 | dpp6 | HbA1c | 0.101 | 431 | nxph3 | ALT | -0.125 |
| 12 | acta2 | Total protein | -0.06 | 222 | dpp6 | HDL | 0.151 | 432 | nxph3 | ALP | 0.093 |
| 13 | acta2 | Urate | 0.124 | 223 | dpp6 | SHBG | 0.135 | 433 | nxph3 | ApoA | 0.118 |
| 14 | acta2 | Urea | 0.261 | 224 | dpp6 | Testosterone | -0.118 | 434 | nxph3 | AST | 0.047 |
| 15 | adm | Albumin | -0.244 | 225 | dpp6 | Total protein | 0.122 | 435 | nxph3 | CRP | -0.124 |
| 16 | adm | ALP | 0.162 | 226 | dpp6 | Triglycerides | -0.097 | 436 | nxph3 | Calcium | 0.051 |
| 17 | adm | CRP | 0.348 | 227 | dpp6 | Urate | -0.155 | 437 | nxph3 | Cholesterol | 0.105 |
| 18 | adm | Calcium | -0.1 | 228 | dpp6 | Vitamin D | 0.073 | 438 | nxph3 | Cystatin C | 0.166 |
| 19 | adm | Creatinine | 0.051 | 229 | eda2r | ALT | -0.082 | 439 | nxph3 | GGT | -0.113 |
| 20 | adm | Cystatin C | 0.566 | 230 | eda2r | Albumin | -0.193 | 440 | nxph3 | Glucose | -0.122 |
| 21 | adm | Direct bilirubin | -0.071 | 231 | eda2r | ALP | 0.108 | 441 | nxph3 | HDL | 0.16 |
| 22 | adm | GGT | 0.122 | 232 | eda2r | ApoB | -0.102 | 442 | nxph3 | LDL | 0.073 |
| 23 | adm | Glucose | 0.119 | 233 | eda2r | CRP | 0.142 | 443 | nxph3 | Phosphate | 0.061 |
| 24 | adm | HbA1c | 0.12 | 234 | eda2r | Calcium | -0.059 | 444 | nxph3 | SHBG | 0.193 |
| 25 | adm | HDL | -0.055 | 235 | eda2r | Cholesterol | -0.099 | 445 | nxph3 | Testosterone | -0.107 |
| 26 | adm | IGF-1 | -0.152 | 236 | eda2r | Creatinine | 0.103 | 446 | nxph3 | Triglycerides | -0.072 |
| 27 | adm | SHBG | -0.05 | 237 | eda2r | Cystatin C | 0.585 | 447 | nxph3 | Urate | -0.057 |
| 28 | adm | Testosterone | -0.091 | 238 | eda2r | Glucose | 0.096 | 448 | nxph3 | Urea | 0.129 |
| 29 | adm | Total bilirubin | -0.12 | 239 | eda2r | HbA1c | 0.181 | 449 | pgf | ALT | 0.088 |
| 30 | adm | Total protein | -0.134 | 240 | eda2r | HDL | -0.055 | 450 | pgf | Albumin | -0.185 |
| 31 | adm | Triglycerides | 0.182 | 241 | eda2r | IGF-1 | -0.099 | 451 | pgf | ALP | 0.119 |
| 32 | adm | Urate | 0.225 | 242 | eda2r | LDL | -0.098 | 452 | pgf | ApoA | -0.232 |
| 33 | adm | Urea | 0.206 | 243 | eda2r | Lipoprotein A | -0.081 | 453 | pgf | ApoB | -0.091 |
| 34 | adm | Vitamin D | -0.139 | 244 | eda2r | SHBG | 0.125 | 454 | pgf | AST | 0.089 |
| 35 | angptl3 | ALT | -0.114 | 245 | eda2r | Urate | 0.157 | 455 | pgf | CRP | 0.194 |
| 36 | angptl3 | Albumin | -0.183 | 246 | eda2r | Urea | 0.256 | 456 | pgf | Calcium | -0.122 |
| 37 | angptl3 | ALP | 0.183 | 247 | efcab14 | Albumin | -0.145 | 457 | pgf | Cholesterol | -0.165 |
| 38 | angptl3 | ApoA | 0.263 | 248 | efcab14 | ALP | 0.175 | 458 | pgf | Creatinine | 0.294 |
| 39 | angptl3 | ApoB | 0.219 | 249 | efcab14 | ApoA | -0.087 | 459 | pgf | Cystatin C | 0.626 |
| 40 | angptl3 | AST | -0.053 | 250 | efcab14 | CRP | 0.16 | 460 | pgf | Direct bilirubin | 0.126 |
| 41 | angptl3 | CRP | 0.196 | 251 | efcab14 | Creatinine | 0.168 | 461 | pgf | GGT | 0.18 |
| 42 | angptl3 | Cholesterol | 0.355 | 252 | efcab14 | Cystatin C | 0.633 | 462 | pgf | Glucose | 0.062 |
| 43 | angptl3 | Creatinine | -0.187 | 253 | efcab14 | GGT | 0.064 | 463 | pgf | HbA1c | 0.088 |
| 44 | angptl3 | Cystatin C | 0.161 | 254 | efcab14 | Glucose | 0.068 | 464 | pgf | HDL | -0.285 |
| 45 | angptl3 | Direct bilirubin | -0.189 | 255 | efcab14 | HbA1c | 0.072 | 465 | pgf | IGF-1 | 0.052 |
| 46 | angptl3 | GGT | -0.069 | 256 | efcab14 | HDL | -0.143 | 466 | pgf | LDL | -0.123 |
| 47 | angptl3 | HDL | 0.295 | 257 | efcab14 | Total protein | -0.08 | 467 | pgf | Lipoprotein A | -0.088 |
| 48 | angptl3 | IGF-1 | -0.097 | 258 | efcab14 | Triglycerides | 0.259 | 468 | pgf | Phosphate | -0.139 |
| 49 | angptl3 | LDL | 0.299 | 259 | efcab14 | Urate | 0.212 | 469 | pgf | SHBG | -0.094 |
| 50 | angptl3 | Oestradiol | 0.163 | 260 | efcab14 | Urea | 0.259 | 470 | pgf | Testosterone | 0.282 |
| 51 | angptl3 | Phosphate | 0.12 | 261 | efemp1 | ALT | -0.076 | 471 | pgf | Total bilirubin | 0.106 |
| 52 | angptl3 | SHBG | 0.204 | 262 | efemp1 | Albumin | -0.33 | 472 | pgf | Triglycerides | 0.194 |
| 53 | angptl3 | Testosterone | -0.375 | 263 | efemp1 | ALP | 0.191 | 473 | pgf | Urate | 0.295 |
| 54 | angptl3 | Total bilirubin | -0.208 | 264 | efemp1 | ApoB | -0.091 | 474 | pgf | Urea | 0.213 |
| 55 | angptl3 | Urate | -0.095 | 265 | efemp1 | CRP | 0.251 | 475 | psap | ALT | 0.052 |
| 56 | angptl3 | Urea | 0.048 | 266 | efemp1 | Calcium | -0.123 | 476 | psap | Albumin | -0.131 |
| 57 | angptl3 | Vitamin D | -0.079 | 267 | efemp1 | Cholesterol | -0.08 | 477 | psap | ALP | 0.221 |
| 58 | b4gat1 | Albumin | 0.111 | 268 | efemp1 | Cystatin C | 0.492 | 478 | psap | ApoA | -0.131 |
| 59 | b4gat1 | ALP | 0.086 | 269 | efemp1 | Glucose | 0.059 | 479 | psap | AST | 0.108 |
| 60 | b4gat1 | ApoA | 0.26 | 270 | efemp1 | HbA1c | 0.176 | 480 | psap | CRP | 0.235 |
| 61 | b4gat1 | ApoB | 0.109 | 271 | efemp1 | IGF-1 | -0.18 | 481 | psap | Creatinine | 0.114 |
| 62 | b4gat1 | AST | 0.158 | 272 | efemp1 | LDL | -0.077 | 482 | psap | Cystatin C | 0.511 |
| 63 | b4gat1 | CRP | -0.067 | 273 | efemp1 | SHBG | 0.142 | 483 | psap | Direct bilirubin | 0.058 |
| 64 | b4gat1 | Calcium | 0.145 | 274 | efemp1 | Testosterone | -0.128 | 484 | psap | GGT | 0.142 |
| 65 | b4gat1 | Cholesterol | 0.201 | 275 | efemp1 | Total bilirubin | -0.106 | 485 | psap | Glucose | 0.063 |
| 66 | b4gat1 | Creatinine | -0.082 | 276 | efemp1 | Triglycerides | -0.087 | 486 | psap | HbA1c | 0.104 |
| 67 | b4gat1 | Direct bilirubin | -0.053 | 277 | efemp1 | Urate | 0.052 | 487 | psap | HDL | -0.167 |
| 68 | b4gat1 | GGT | -0.058 | 278 | efemp1 | Urea | 0.145 | 488 | psap | IGF-1 | -0.083 |
| 69 | b4gat1 | HbA1c | 0.049 | 279 | efhd1 | ALT | 0.107 | 489 | psap | Testosterone | 0.051 |
| 70 | b4gat1 | HDL | 0.27 | 280 | efhd1 | Albumin | -0.113 | 490 | psap | Total protein | 0.06 |
| 71 | b4gat1 | LDL | 0.141 | 281 | efhd1 | AST | 0.136 | 491 | psap | Triglycerides | 0.139 |
| 72 | b4gat1 | Lipoprotein A | 0.056 | 282 | efhd1 | CRP | 0.107 | 492 | psap | Urate | 0.186 |
| 73 | b4gat1 | Phosphate | 0.114 | 283 | efhd1 | Calcium | -0.078 | 493 | psap | Urea | 0.127 |
| 74 | b4gat1 | SHBG | 0.073 | 284 | efhd1 | Creatinine | 0.198 | 494 | psap | Vitamin D | -0.051 |
| 75 | b4gat1 | Testosterone | -0.195 | 285 | efhd1 | Cystatin C | 0.425 | 495 | psg1 | ALT | -0.121 |
| 76 | b4gat1 | Total protein | 0.137 | 286 | efhd1 | GGT | 0.137 | 496 | psg1 | Albumin | -0.081 |
| 77 | b4gat1 | Triglycerides | -0.072 | 287 | efhd1 | HbA1c | 0.061 | 497 | psg1 | ApoA | 0.136 |
| 78 | b4gat1 | Urate | -0.055 | 288 | efhd1 | Phosphate | -0.09 | 498 | psg1 | ApoB | -0.05 |
| 79 | bcan | ALT | -0.069 | 289 | efhd1 | Testosterone | 0.092 | 499 | psg1 | Creatinine | -0.057 |
| 80 | bcan | Albumin | 0.081 | 290 | efhd1 | Total bilirubin | 0.052 | 500 | psg1 | Cystatin C | 0.076 |
| 81 | bcan | ApoA | 0.142 | 291 | efhd1 | Total protein | -0.111 | 501 | psg1 | GGT | -0.121 |
| 82 | bcan | ApoB | 0.081 | 292 | efhd1 | Triglycerides | 0.082 | 502 | psg1 | Glucose | 0.067 |
| 83 | bcan | AST | 0.11 | 293 | efhd1 | Urate | 0.185 | 503 | psg1 | HDL | 0.163 |
| 84 | bcan | CRP | -0.13 | 294 | efhd1 | Urea | 0.281 | 504 | psg1 | IGF-1 | -0.122 |
| 85 | bcan | Calcium | 0.091 | 295 | igfbp4 | ALT | -0.047 | 505 | psg1 | SHBG | 0.22 |
| 86 | bcan | Cholesterol | 0.142 | 296 | igfbp4 | Albumin | -0.198 | 506 | psg1 | Testosterone | -0.118 |
| 87 | bcan | Creatinine | -0.059 | 297 | igfbp4 | ALP | 0.168 | 507 | psg1 | Total protein | -0.068 |
| 88 | bcan | Cystatin C | -0.049 | 298 | igfbp4 | ApoA | -0.08 | 508 | psg1 | Triglycerides | -0.079 |
| 89 | bcan | GGT | -0.121 | 299 | igfbp4 | ApoB | -0.053 | 509 | psg1 | Urate | -0.095 |
| 90 | bcan | Glucose | -0.126 | 300 | igfbp4 | CRP | 0.271 | 510 | psg1 | Urea | 0.099 |
| 91 | bcan | HbA1c | -0.069 | 301 | igfbp4 | Calcium | -0.05 | 511 | reg1a | ALT | -0.087 |
| 92 | bcan | HDL | 0.177 | 302 | igfbp4 | Cholesterol | -0.082 | 512 | reg1a | Albumin | -0.112 |
| 93 | bcan | LDL | 0.122 | 303 | igfbp4 | Creatinine | 0.211 | 513 | reg1a | ALP | 0.1 |
| 94 | bcan | SHBG | 0.123 | 304 | igfbp4 | Cystatin C | 0.707 | 514 | reg1a | ApoB | -0.08 |
| 95 | bcan | Testosterone | -0.121 | 305 | igfbp4 | GGT | 0.121 | 515 | reg1a | AST | -0.059 |
| 96 | bcan | Triglycerides | -0.125 | 306 | igfbp4 | Glucose | 0.095 | 516 | reg1a | CRP | 0.052 |
| 97 | bcan | Urate | -0.148 | 307 | igfbp4 | HbA1c | 0.151 | 517 | reg1a | Cholesterol | -0.077 |
| 98 | bcan | Vitamin D | 0.111 | 308 | igfbp4 | HDL | -0.137 | 518 | reg1a | Creatinine | 0.137 |
| 99 | cd7 | ALT | -0.079 | 309 | igfbp4 | IGF-1 | -0.057 | 519 | reg1a | Cystatin C | 0.407 |
| 100 | cd7 | Albumin | -0.16 | 310 | igfbp4 | LDL | -0.076 | 520 | reg1a | Glucose | 0.061 |
| 101 | cd7 | ALP | 0.162 | 311 | igfbp4 | Phosphate | -0.061 | 521 | reg1a | HbA1c | 0.108 |
| 102 | cd7 | ApoA | -0.062 | 312 | igfbp4 | SHBG | -0.068 | 522 | reg1a | LDL | -0.075 |
| 103 | cd7 | AST | -0.07 | 313 | igfbp4 | Testosterone | 0.09 | 523 | reg1a | SHBG | 0.058 |
| 104 | cd7 | CRP | 0.158 | 314 | igfbp4 | Triglycerides | 0.163 | 524 | reg1a | Total bilirubin | -0.059 |
| 105 | cd7 | Cystatin C | 0.356 | 315 | igfbp4 | Urate | 0.297 | 525 | reg1a | Urate | 0.139 |
| 106 | cd7 | Direct bilirubin | -0.09 | 316 | igfbp4 | Urea | 0.268 | 526 | reg1a | Urea | 0.242 |
| 107 | cd7 | HbA1c | 0.053 | 317 | igfbp4 | Vitamin D | -0.056 | 527 | ret | ALT | 0.242 |
| 108 | cd7 | HDL | -0.075 | 318 | iglc2 | Albumin | -0.264 | 528 | ret | Albumin | 0.142 |
| 109 | cd7 | Oestradiol | 0.181 | 319 | iglc2 | ALP | 0.092 | 529 | ret | ALP | 0.121 |
| 110 | cd7 | Phosphate | 0.066 | 320 | iglc2 | ApoA | -0.191 | 530 | ret | ApoA | -0.071 |
| 111 | cd7 | SHBG | 0.053 | 321 | iglc2 | ApoB | -0.103 | 531 | ret | ApoB | 0.075 |
| 112 | cd7 | Testosterone | -0.121 | 322 | iglc2 | CRP | 0.21 | 532 | ret | AST | 0.213 |
| 113 | cd7 | Total bilirubin | -0.137 | 323 | iglc2 | Calcium | -0.107 | 533 | ret | CRP | 0.111 |
| 114 | cd7 | Urea | 0.108 | 324 | iglc2 | Cholesterol | -0.146 | 534 | ret | Calcium | 0.152 |
| 115 | cd99l2 | Albumin | -0.115 | 325 | iglc2 | Creatinine | 0.139 | 535 | ret | GGT | 0.18 |
| 116 | cd99l2 | ALP | 0.23 | 326 | iglc2 | Cystatin C | 0.379 | 536 | ret | Glucose | 0.094 |
| 117 | cd99l2 | ApoA | 0.158 | 327 | iglc2 | GGT | 0.083 | 537 | ret | HbA1c | 0.179 |
| 118 | cd99l2 | ApoB | 0.095 | 328 | iglc2 | HbA1c | 0.095 | 538 | ret | HDL | -0.128 |
| 119 | cd99l2 | CRP | 0.293 | 329 | iglc2 | HDL | -0.2 | 539 | ret | LDL | 0.05 |
| 120 | cd99l2 | Cholesterol | 0.16 | 330 | iglc2 | IGF-1 | -0.056 | 540 | ret | SHBG | -0.233 |
| 121 | cd99l2 | Cystatin C | 0.431 | 331 | iglc2 | LDL | -0.12 | 541 | ret | Testosterone | -0.087 |
| 122 | cd99l2 | GGT | 0.085 | 332 | iglc2 | Phosphate | -0.061 | 542 | ret | Total protein | 0.155 |
| 123 | cd99l2 | Glucose | 0.078 | 333 | iglc2 | Testosterone | 0.092 | 543 | ret | Triglycerides | 0.222 |
| 124 | cd99l2 | HbA1c | 0.077 | 334 | iglc2 | Total bilirubin | -0.095 | 544 | ret | Urate | 0.082 |
| 125 | cd99l2 | HDL | 0.108 | 335 | iglc2 | Total protein | 0.173 | 545 | ret | Urea | -0.084 |
| 126 | cd99l2 | IGF-1 | -0.087 | 336 | iglc2 | Triglycerides | 0.066 | 546 | ret | Vitamin D | -0.067 |
| 127 | cd99l2 | LDL | 0.125 | 337 | iglc2 | Urate | 0.191 | 547 | shisa5 | Albumin | -0.243 |
| 128 | cd99l2 | SHBG | -0.072 | 338 | iglc2 | Urea | 0.122 | 548 | shisa5 | ALP | 0.198 |
| 129 | cd99l2 | Testosterone | -0.089 | 339 | mmp12 | ALT | -0.057 | 549 | shisa5 | ApoA | -0.148 |
| 130 | cd99l2 | Total bilirubin | -0.074 | 340 | mmp12 | Albumin | -0.085 | 550 | shisa5 | ApoB | -0.063 |
| 131 | cd99l2 | Triglycerides | 0.155 | 341 | mmp12 | ALP | 0.145 | 551 | shisa5 | CRP | 0.317 |
| 132 | cd99l2 | Urate | 0.146 | 342 | mmp12 | ApoA | -0.111 | 552 | shisa5 | Calcium | -0.099 |
| 133 | cd99l2 | Urea | 0.125 | 343 | mmp12 | CRP | 0.203 | 553 | shisa5 | Cholesterol | -0.103 |
| 134 | cd99l2 | Vitamin D | -0.132 | 344 | mmp12 | Cystatin C | 0.305 | 554 | shisa5 | Creatinine | 0.168 |
| 135 | cdsn | ALT | -0.049 | 345 | mmp12 | Direct bilirubin | -0.071 | 555 | shisa5 | Cystatin C | 0.746 |
| 136 | cdsn | Albumin | -0.065 | 346 | mmp12 | GGT | 0.049 | 556 | shisa5 | GGT | 0.088 |
| 137 | cdsn | Calcium | -0.057 | 347 | mmp12 | HbA1c | 0.138 | 557 | shisa5 | Glucose | 0.082 |
| 138 | cdsn | Creatinine | 0.187 | 348 | mmp12 | HDL | -0.119 | 558 | shisa5 | HbA1c | 0.152 |
| 139 | cdsn | Cystatin C | 0.301 | 349 | mmp12 | IGF-1 | -0.15 | 559 | shisa5 | HDL | -0.178 |
| 140 | cdsn | Phosphate | -0.057 | 350 | mmp12 | Oestradiol | -0.177 | 560 | shisa5 | IGF-1 | -0.081 |
| 141 | cdsn | Testosterone | 0.096 | 351 | mmp12 | Phosphate | 0.051 | 561 | shisa5 | LDL | -0.072 |
| 142 | cdsn | Total protein | -0.075 | 352 | mmp12 | SHBG | 0.067 | 562 | shisa5 | Lipoprotein A | -0.061 |
| 143 | cdsn | Urate | 0.162 | 353 | mmp12 | Total bilirubin | -0.097 | 563 | shisa5 | Testosterone | 0.06 |
| 144 | cdsn | Urea | 0.151 | 354 | mmp12 | Total protein | 0.086 | 564 | shisa5 | Triglycerides | 0.116 |
| 145 | cdsn | Vitamin D | 0.062 | 355 | mmp12 | Triglycerides | 0.08 | 565 | shisa5 | Urate | 0.245 |
| 146 | chga | ALT | -0.123 | 356 | mmp12 | Urate | 0.103 | 566 | shisa5 | Urea | 0.253 |
| 147 | chga | Albumin | -0.14 | 357 | mmp12 | Urea | 0.079 | 567 | shisa5 | Vitamin D | -0.074 |
| 148 | chga | ALP | 0.093 | 358 | mmp3 | ALT | 0.064 | 568 | tff2 | ALT | -0.065 |
| 149 | chga | ApoB | -0.153 | 359 | mmp3 | ALP | -0.131 | 569 | tff2 | Albumin | -0.115 |
| 150 | chga | Calcium | -0.059 | 360 | mmp3 | ApoA | -0.178 | 570 | tff2 | ALP | 0.11 |
| 151 | chga | Cholesterol | -0.112 | 361 | mmp3 | ApoB | -0.124 | 571 | tff2 | ApoB | -0.076 |
| 152 | chga | Creatinine | 0.099 | 362 | mmp3 | AST | 0.057 | 572 | tff2 | CRP | 0.056 |
| 153 | chga | Cystatin C | 0.31 | 363 | mmp3 | Calcium | -0.103 | 573 | tff2 | Cholesterol | -0.055 |
| 154 | chga | Direct bilirubin | -0.075 | 364 | mmp3 | Cholesterol | -0.171 | 574 | tff2 | Creatinine | 0.061 |
| 155 | chga | Glucose | 0.075 | 365 | mmp3 | Creatinine | 0.495 | 575 | tff2 | Cystatin C | 0.346 |
| 156 | chga | HbA1c | 0.115 | 366 | mmp3 | Cystatin C | 0.283 | 576 | tff2 | Direct bilirubin | -0.075 |
| 157 | chga | HDL | 0.062 | 367 | mmp3 | Direct bilirubin | 0.225 | 577 | tff2 | Glucose | 0.084 |
| 158 | chga | LDL | -0.124 | 368 | mmp3 | GGT | 0.146 | 578 | tff2 | HbA1c | 0.153 |
| 159 | chga | SHBG | 0.087 | 369 | mmp3 | HDL | -0.16 | 579 | tff2 | LDL | -0.069 |
| 160 | chga | Total bilirubin | -0.141 | 370 | mmp3 | IGF-1 | 0.151 | 580 | tff2 | SHBG | 0.062 |
| 161 | chga | Total protein | -0.068 | 371 | mmp3 | LDL | -0.143 | 581 | tff2 | Testosterone | -0.102 |
| 162 | chga | Triglycerides | -0.105 | 372 | mmp3 | Lipoprotein A | -0.057 | 582 | tff2 | Total bilirubin | -0.122 |
| 163 | chga | Urea | 0.232 | 373 | mmp3 | Oestradiol | -0.263 | 583 | tff2 | Urate | 0.063 |
| 164 | clec14a | Albumin | -0.127 | 374 | mmp3 | Phosphate | -0.238 | 584 | tff2 | Urea | 0.191 |
| 165 | clec14a | ALP | 0.141 | 375 | mmp3 | SHBG | -0.142 | 585 | tmprss5 | ALT | -0.079 |
| 166 | clec14a | ApoA | -0.084 | 376 | mmp3 | Testosterone | 0.588 | 586 | tmprss5 | ALP | 0.05 |
| 167 | clec14a | ApoB | -0.111 | 377 | mmp3 | Total bilirubin | 0.189 | 587 | tmprss5 | ApoA | 0.063 |
| 168 | clec14a | AST | 0.086 | 378 | mmp3 | Total protein | -0.072 | 588 | tmprss5 | AST | 0.05 |
| 169 | clec14a | CRP | 0.068 | 379 | mmp3 | Urate | 0.291 | 589 | tmprss5 | CRP | -0.135 |
| 170 | clec14a | Calcium | -0.065 | 380 | mmp3 | Urea | 0.2 | 590 | tmprss5 | Cholesterol | 0.086 |
| 171 | clec14a | Cholesterol | -0.11 | 381 | msln | ALT | -0.076 | 591 | tmprss5 | GGT | -0.085 |
| 172 | clec14a | Creatinine | 0.222 | 382 | msln | Albumin | -0.141 | 592 | tmprss5 | HDL | 0.11 |
| 173 | clec14a | Cystatin C | 0.518 | 383 | msln | ALP | 0.069 | 593 | tmprss5 | IGF-1 | 0.068 |
| 174 | clec14a | Direct bilirubin | 0.074 | 384 | msln | AST | -0.054 | 594 | tmprss5 | LDL | 0.067 |
| 175 | clec14a | HbA1c | 0.073 | 385 | msln | CRP | 0.09 | 595 | tmprss5 | SHBG | 0.096 |
| 176 | clec14a | HDL | -0.09 | 386 | msln | Creatinine | 0.08 | 596 | tmprss5 | Testosterone | -0.103 |
| 177 | clec14a | LDL | -0.109 | 387 | msln | Cystatin C | 0.245 | 597 | tmprss5 | Triglycerides | -0.075 |
| 178 | clec14a | Lipoprotein A | -0.068 | 388 | msln | HbA1c | 0.051 | 598 | tmprss5 | Urate | -0.139 |
| 179 | clec14a | SHBG | 0.055 | 389 | msln | SHBG | 0.076 | 599 | tnc | ALT | -0.085 |
| 180 | clec14a | Testosterone | 0.108 | 390 | msln | Total protein | -0.073 | 600 | tnc | Albumin | -0.146 |
| 181 | clec14a | Total protein | -0.099 | 391 | msln | Urate | 0.056 | 601 | tnc | ALP | 0.201 |
| 182 | clec14a | Triglycerides | 0.048 | 392 | msln | Urea | 0.096 | 602 | tnc | CRP | 0.154 |
| 183 | clec14a | Urate | 0.158 | 393 | nppb | ALT | -0.101 | 603 | tnc | Cystatin C | 0.121 |
| 184 | clec14a | Urea | 0.219 | 394 | nppb | Albumin | -0.131 | 604 | tnc | HbA1c | -0.074 |
| 185 | cthrc1 | ALT | 0.147 | 395 | nppb | ApoA | 0.056 | 605 | tnc | HDL | 0.081 |
| 186 | cthrc1 | Albumin | -0.163 | 396 | nppb | ApoB | -0.099 | 606 | tnc | SHBG | 0.138 |
| 187 | cthrc1 | ALP | 0.105 | 397 | nppb | Calcium | -0.068 | 607 | tnc | Testosterone | -0.063 |
| 188 | cthrc1 | ApoA | -0.126 | 398 | nppb | Cholesterol | -0.072 | 608 | tnc | Triglycerides | -0.08 |
| 189 | cthrc1 | ApoB | -0.047 | 399 | nppb | Creatinine | -0.051 | 609 | tnc | Urea | 0.06 |
| 190 | cthrc1 | AST | 0.181 | 400 | nppb | Cystatin C | 0.114 | 610 | tnfrsf10b | Albumin | -0.218 |
| 191 | cthrc1 | CRP | 0.162 | 401 | nppb | GGT | -0.059 | 611 | tnfrsf10b | ALP | 0.178 |
| 192 | cthrc1 | Calcium | -0.135 | 402 | nppb | HDL | 0.076 | 612 | tnfrsf10b | ApoA | -0.149 |
| 193 | cthrc1 | Cholesterol | -0.083 | 403 | nppb | IGF-1 | -0.116 | 613 | tnfrsf10b | ApoB | -0.082 |
| 194 | cthrc1 | Creatinine | 0.204 | 404 | nppb | LDL | -0.088 | 614 | tnfrsf10b | AST | 0.054 |
| 195 | cthrc1 | Cystatin C | 0.284 | 405 | nppb | Phosphate | 0.069 | 615 | tnfrsf10b | CRP | 0.269 |
| 196 | cthrc1 | Direct bilirubin | 0.083 | 406 | nppb | SHBG | 0.198 | 616 | tnfrsf10b | Calcium | -0.063 |
| 197 | cthrc1 | GGT | 0.192 | 407 | nppb | Testosterone | -0.063 | 617 | tnfrsf10b | Cholesterol | -0.121 |
| 198 | cthrc1 | Glucose | 0.09 | 408 | nppb | Total protein | -0.109 | 618 | tnfrsf10b | Creatinine | 0.131 |
| 199 | cthrc1 | HbA1c | 0.134 | 409 | nppb | Triglycerides | -0.118 | 619 | tnfrsf10b | Cystatin C | 0.651 |
| 200 | cthrc1 | HDL | -0.144 | 410 | nppb | Urea | 0.125 | 620 | tnfrsf10b | GGT | 0.145 |
| 201 | cthrc1 | IGF-1 | -0.131 | 411 | nppb | Vitamin D | 0.055 | 621 | tnfrsf10b | Glucose | 0.085 |
| 202 | cthrc1 | LDL | -0.069 | 412 | ntprobnp | ALT | -0.167 | 622 | tnfrsf10b | HbA1c | 0.197 |
| 203 | cthrc1 | Oestradiol | -0.231 | 413 | ntprobnp | Albumin | -0.182 | 623 | tnfrsf10b | HDL | -0.207 |
| 204 | cthrc1 | Phosphate | -0.097 | 414 | ntprobnp | ApoA | 0.078 | 624 | tnfrsf10b | IGF-1 | -0.116 |
| 205 | cthrc1 | SHBG | -0.093 | 415 | ntprobnp | ApoB | -0.115 | 625 | tnfrsf10b | LDL | -0.105 |
| 206 | cthrc1 | Testosterone | 0.246 | 416 | ntprobnp | Calcium | -0.072 | 626 | tnfrsf10b | Total bilirubin | -0.066 |
| 207 | cthrc1 | Total protein | -0.051 | 417 | ntprobnp | Cholesterol | -0.072 | 627 | tnfrsf10b | Triglycerides | 0.198 |
| 208 | cthrc1 | Triglycerides | 0.127 | 418 | ntprobnp | Creatinine | -0.057 | 628 | tnfrsf10b | Urate | 0.238 |
| 209 | cthrc1 | Urate | 0.213 | 419 | ntprobnp | Cystatin C | 0.212 | 629 | tnfrsf10b | Urea | 0.206 |
| 210 | cthrc1 | Urea | 0.128 | 420 | ntprobnp | GGT | -0.091 | 630 | tnfrsf10b | Vitamin D | -0.082 |

**Supplementary Table 5 | Two-sample Mendelian Randomization results linking protein levels to cardiovascular outcomes and CKD.** We queried the Proteome Phenome Atlas to identify evidence of causal relationships between genetically predicted protein levels and cardiovascular or kidney outcomes using two-sample MR. The table includes effect estimates, standard errors, and significant values for each protein-outcome pair. Results support potential causal roles for selected proteins in CVE and CKD pathophysiology.

| **Chronic Kideney Disease --> Proteins** | | | | | | | |
| --- | --- | --- | --- | --- | --- | --- | --- |
| **Disease** |  | **Protein** | **Protein_definition** | **method** | **OR [95% CI]** | **P value** | **QTL_type** |
| N14_CHRONKIDNEYDIS | Chronic Kidendy Disease | CLEC14A | C-type lectin domain family 14 member A | Wald ratio | 1.33 [1.23-1.43] | 3.96E-14 | trans |
| N14_CHRONKIDNEYDIS | Chronic Kidendy Disease | IGFBP4 | Insulin-like growth factor-binding protein 4 | Wald ratio | 1.32 [1.22-1.42] | 1.76E-13 | trans |
| N14_CHRONKIDNEYDIS | Chronic Kidendy Disease | SHISA5 | Protein shisa-5 | Wald ratio | 1.31 [1.22-1.41] | 3.01E-13 | trans |
| N14_CHRONKIDNEYDIS | Chronic Kidendy Disease | CDSN | Corneodesmosin | Wald ratio | 1.26 [1.18-1.34] | 6.77E-12 | trans |
| N14_CHRONKIDNEYDIS | Chronic Kidendy Disease | REG1A | Lithostathine-1-alpha | Wald ratio | 1.28 [1.19-1.38] | 2.48E-11 | trans |
| N14_CHRONKIDNEYDIS | Chronic Kidendy Disease | EDA2R | Tumor necrosis factor receptor superfamily member 27 | Wald ratio | 1.2 [1.13-1.27] | 5.06E-09 | trans |
| N14_CHRONKIDNEYDIS | Chronic Kidendy Disease | CHGA | Chromogranin-A | Wald ratio | 1.25 [1.16-1.34] | 1.05E-08 | trans |
| N14_CHRONKIDNEYDIS | Chronic Kidendy Disease | TNFRSF10B | Tumor necrosis factor receptor superfamily member 10B | Wald ratio | 1.21 [1.13-1.3] | 3.57E-08 | trans |
| N14_CHRONKIDNEYDIS | Chronic Kidendy Disease | EFEMP1 | EGF-containing fibulin-like extracellular matrix protein 1 | Wald ratio | 1.22 [1.13-1.31] | 3.97E-08 | trans |
| N14_CHRONKIDNEYDIS | Chronic Kidendy Disease | EFCAB14 | EF-hand calcium-binding domain-containing protein 14 | Wald ratio | 1.23 [1.14-1.33] | 5.20E-08 | trans |
| N14_CHRONKIDNEYDIS | Chronic Kidendy Disease | CD7 | T-cell antigen CD7 | Wald ratio | 1.22 [1.13-1.31] | 1.20E-07 | trans |
| N14_CHRONKIDNEYDIS | Chronic Kidendy Disease | ADM | Pro-adrenomedullin | Wald ratio | 1.21 [1.13-1.31] | 1.30E-07 | trans |
| N14_CHRONKIDNEYDIS | Chronic Kidendy Disease | MMP3 | Stromelysin-1 | Wald ratio | 1.14 [1.08-1.21] | 1.51E-06 | trans |
| N14_CHRONKIDNEYDIS | Chronic Kidendy Disease | NXPH3 | Neurexophilin-3 | Wald ratio | 1.2 [1.11-1.3] | 1.76E-06 | trans |
| N14_CHRONKIDNEYDIS | Chronic Kidendy Disease | EFHD1 | EF-hand domain-containing protein D1 | Wald ratio | 1.19 [1.11-1.28] | 2.15E-06 | trans |
| N14_CHRONKIDNEYDIS | Chronic Kidendy Disease | PGF | Placenta growth factor | Wald ratio | 1.17 [1.09-1.25] | 1.21E-05 | trans |
| N14_CHRONKIDNEYDIS | Chronic Kidendy Disease | ACTA2 | Actin, aortic smooth muscle | Wald ratio | 1.16 [1.08-1.24] | 5.55E-05 | trans |
| N14_CHRONKIDNEYDIS | Chronic Kidendy Disease | TFF2 | Trefoil factor 2 | Wald ratio | 1.16 [1.08-1.25] | 8.43E-05 | trans |
| N14_CHRONKIDNEYDIS | Chronic Kidendy Disease | PSAP | Prosaposin | Wald ratio | 1.16 [1.07-1.25] | 0.000112273 | trans |
| N14_CHRONKIDNEYDIS | Chronic Kidendy Disease | CD99L2 | CD99 antigen-like protein 2 | Wald ratio | 1.13 [1.05-1.22] | 0.001563039 | trans |
| N14_CHRONKIDNEYDIS | Chronic Kidendy Disease | IGLC2 | Immunoglobulin lambda constant 2 | Wald ratio | 1.11 [1.03-1.2] | 0.005801815 | trans |
| N14_CHRONKIDNEYDIS | Chronic Kidendy Disease | ANGPTL3 | Angiopoietin-related protein 3 | Wald ratio | 1.09 [1.02-1.17] | 0.012210875 | trans |
| N14_CHRONKIDNEYDIS | Chronic Kidendy Disease | B4GAT1 | Beta-1,4-glucuronyltransferase 1 | Wald ratio | 1.09 [1.01-1.17] | 0.026392334 | trans |
| N14_CHRONKIDNEYDIS | Chronic Kidendy Disease | MMP12 | Macrophage metalloelastase | Wald ratio | 1.07 [1.01-1.15] | 0.03464918 | trans |
| N14_CHRONKIDNEYDIS | Chronic Kidendy Disease | MSLN | Mesothelin | Wald ratio | 1.08 [1-1.17] | 0.041305035 | trans |
| **Protein --> Cardiovascular Events** | | | | | | | |
| **Disease** |  | **Protein** | **Protein_definition** | **method** | **OR [95% CI]** | **P value** | **QTL_type** |
| I9_CORATHER | coronary atherosclerosis | NTproBNP | N-terminal prohormone of brain natriuretic peptide | Inverse variance weighted | 0.92 [0.87-0.97] | 0.001685427 | cis |
| I9_CORATHER | coronary atherosclerosis | NPPB | Natriuretic peptides B | Inverse variance weighted | 0.89 [0.8-0.98] | 0.021286652 | cis |
| I9_CORATHER | coronary atherosclerosis | ANGPTL3 | Angiopoietin-related protein 3 | Inverse variance weighted | 0.51 [0.42-0.64] | 7.19E-10 | trans |
| I9_CORATHER | coronary atherosclerosis | MSLN | Mesothelin | Inverse variance weighted | 1.18 [1.05-1.33] | 0.004993563 | trans |
| I9_CORATHER | coronary atherosclerosis | MMP3 | Stromelysin-1 | Inverse variance weighted | 1.37 [1.03-1.81] | 0.031853935 | trans |
| I9_K_CARDIAC | Death due to cardiac causes | MMP3 | Stromelysin-1 | Inverse variance weighted | 0.94 [0.89-0.98] | 0.00987345 | cis |
| I9_K_CARDIAC | Death due to cardiac causes | NTproBNP | N-terminal prohormone of brain natriuretic peptide | Inverse variance weighted | 1.08 [1.01-1.17] | 0.029450709 | cis |
| I9_K_CARDIAC | Death due to cardiac causes | NPPB | Natriuretic peptides B | Inverse variance weighted | 1.13 [1-1.28] | 0.043632128 | cis |
| I9_K_CARDIAC | Death due to cardiac causes | TNFRSF10B | Tumor necrosis factor receptor superfamily member 10B | Inverse variance weighted | 0.72 [0.57-0.9] | 0.004392774 | trans |
| I9_HEARTFAIL | Heart Failure | EFCAB14 | EF-hand calcium-binding domain-containing protein 14 | Inverse variance weighted | 0.82 [0.67-1] | 0.050491082 | cis |
| I9_HEARTFAIL | Heart Failure | TNFRSF10B | Tumor necrosis factor receptor superfamily member 10B | Inverse variance weighted | 0.83 [0.68-1.01] | 0.058485089 | trans |
| I9_IHD | Ischaemic heart disease | NTproBNP | N-terminal prohormone of brain natriuretic peptide | Inverse variance weighted | 0.93 [0.89-0.97] | 0.001860333 | cis |
| I9_IHD | Ischaemic heart disease | EFEMP1 | EGF-containing fibulin-like extracellular matrix protein 1 | Inverse variance weighted | 1.08 [1.02-1.16] | 0.012572526 | cis |
| I9_IHD | Ischaemic heart disease | NPPB | Natriuretic peptides B | Inverse variance weighted | 0.9 [0.83-0.98] | 0.012986132 | cis |
| I9_IHD | Ischaemic heart disease | ANGPTL3 | Angiopoietin-related protein 3 | Inverse variance weighted | 0.59 [0.49-0.7] | 1.50E-08 | trans |
| I9_IHD | Ischaemic heart disease | MSLN | Mesothelin | Inverse variance weighted | 1.17 [1.05-1.3] | 0.004547064 | trans |
| I9_IHD | Ischaemic heart disease | MMP3 | Stromelysin-1 | Inverse variance weighted | 1.45 [1.09-1.93] | 0.011504613 | trans |
| I9_IHD | Ischaemic heart disease | MMP12 | Macrophage metalloelastase | Inverse variance weighted | 1.33 [1.05-1.69] | 0.019548116 | trans |
| I9_MI_STRICT | Myocardial infarction | REG1A | Lithostathine-1-alpha | Inverse variance weighted | 1.07 [1.01-1.14] | 0.022398422 | cis |
| I9_MI_STRICT | Myocardial infarction | ANGPTL3 | Angiopoietin-related protein 3 | Inverse variance weighted | 0.56 [0.45-0.69] | 7.20E-08 | trans |
| I9_MI_STRICT | Myocardial infarction | MSLN | Mesothelin | Inverse variance weighted | 1.24 [1.04-1.49] | 0.016076122 | trans |
| I9_STR | Stroke | MMP12 | Macrophage metalloelastase | Inverse variance weighted | 0.94 [0.9-0.98] | 0.001300327 | cis |
| I9_STR | Stroke | TFF2 | Trefoil factor 2 | Inverse variance weighted | 0.89 [0.79-1] | 0.047633769 | cis |
| I9_STR | Stroke | PGF | Placenta growth factor | Inverse variance weighted | 0.78 [0.66-0.92] | 0.004046135 | trans |
| I9_STR | Stroke | REG1A | Lithostathine-1-alpha | Inverse variance weighted | 0.91 [0.85-0.97] | 0.005291886 | trans |
| I9_STR | Stroke | EFEMP1 | EGF-containing fibulin-like extracellular matrix protein 1 | Inverse variance weighted | 1.32 [1.03-1.7] | 0.030777538 | trans |
| I9_CHD | coronary heart disease | EFEMP1 | EGF-containing fibulin-like extracellular matrix protein 1 | Inverse variance weighted | 1.11 [1.04-1.2] | 0.003579229 | cis |
| I9_CHD | coronary heart disease | NTproBNP | N-terminal prohormone of brain natriuretic peptide | Inverse variance weighted | 0.93 [0.88-0.98] | 0.004553238 | cis |
| I9_CHD | coronary heart disease | REG1A | Lithostathine-1-alpha | Inverse variance weighted | 1.06 [1.01-1.11] | 0.011592342 | cis |
| I9_CHD | coronary heart disease | NPPB | Natriuretic peptides B | Inverse variance weighted | 0.9 [0.82-0.98] | 0.013805466 | cis |
| I9_CHD | coronary heart disease | IGLC2 | Immunoglobulin lambda constant 2 | Inverse variance weighted | 0.89 [0.8-0.98] | 0.023266526 | cis |
| I9_CHD | coronary heart disease | ANGPTL3 | Angiopoietin-related protein 3 | Inverse variance weighted | 0.52 [0.45-0.59] | 3.05E-21 | trans |
| I9_AF | Atrial fibrillation and flutter | EFEMP1 | EGF-containing fibulin-like extracellular matrix protein 1 | Inverse variance weighted | 0.84 [0.76-0.94] | 0.002134637 | cis |
| I9_AF | Atrial fibrillation and flutter | EDA2R | Tumor necrosis factor receptor superfamily member 27 | Inverse variance weighted | 0.68 [0.53-0.87] | 0.001883595 | trans |

**Supplementary Table 6 | Tissue-specific expression of the 34 proteins associated with incident CVEs in CKD.** Tissue-level expression patterns were derived from GTEx RNA-seq data by identifying the tissue with the highest expression (maximum log₂(TPM + 1)) for each protein. This analysis highlights the biological relevance of selected proteins, many of which exhibit enriched expression in cardiovascular, vascular, renal, hepatic, or immune-related tissues.

| **Protein** | **Top Tissue** | **Max log2(TPM+1)** | **Protein** | **Top Tissue** | **Max log2(TPM+1)** |
| --- | --- | --- | --- | --- | --- |
| **NPPB** | Heart_Atrial_Appendage | 10.693496 | **B4GAT1** | Brain_Frontal_Cortex_BA9 | 8.103818 |
| **EFCAB14** | Artery_Tibial | 6.046845 | **MMP3** | Minor_Salivary_Gland | 7.449421 |
| **ANGPTL3** | Liver | 7.007835 | **MMP12** | Small_Intestine_Terminal_Ileum_Lymphode_Aggregate | 3.981067 |
| **BCAN** | Brain_Amygdala | 7.584624 | **TMPRSS5** | Nerve_Tibial | 6.476839 |
| **EFEMP1** | Artery_Aorta | 10.621969 | **CLEC14A** | Adipose_Subcutaneous | 7.063298 |
| **REG1A** | Pancreas_Mixed_Cell | 16.375525 | **PGF** | Thyroid | 6.663871 |
| **EFHD1** | Artery_Tibial | 8.612905 | **CHGA** | Pancreas_Islets | 11.263046 |
| **SHISA5** | Whole_Blood | 7.378095 | **MSLN** | Adipose_Visceral_Omentum | 6.428665 |
| **CDSN** | Skin_Sun_Exposed_Lower_leg | 6.733571 | **IGFBP4** | Ovary | 11.344851 |
| **DPP6** | Brain_Cerebellar_Hemisphere | 6.348383 | **NXPH3** | Artery_Aorta | 6.424800 |
| **TNFRSF10B** | Cells_EBV-transformed_lymphocytes | 6.999211 | **CD7** | Whole_Blood | 6.241070 |
| **CTHRC1** | Cells_Cultured_fibroblasts | 7.982851 | **PSG1** | Cells_Cultured_fibroblasts | 0.751105 |
| **TNC** | Artery_Tibial | 8.481840 | **TFF2** | Stomach_Mixed_Cell | 11.622660 |
| **RET** | Brain_Substantia_nigra | 2.522078 | **IGLC2** | Spleen | 11.030039 |
| **PSAP** | Artery_Aorta | 10.732948 | **EDA2R** | Cells_Cultured_fibroblasts | 5.170951 |
| **ACTA2** | Artery_Tibial | 12.630768 | **CD99L2** | Nerve_Tibial | 6.807870 |
| **ADM** | Cells_Cultured_fibroblasts | 9.808570 |  |  |  |

**Supplementary Table 7 | Predictive performance of risk models for CVE in CKD.** Discrimination and predictive values were evaluated across models using the C-index, positive predictive value (PPV), and negative predictive value (NPV) from 1 to 10 years. Models included protein-based risk scores, polygenic risk scores (PRS), Pooled Cohort Equation (PCE), and combinations with eGFR and demographic covariates. The protein-only model showed the highest discrimination (C-index = 0.729) and achieved superior PPV across all time points compared to clinical and genetic models

| **Model** | **Proteins-only** | **Baseline (Age, Sex, Ethnicity, eGFR)** | **PCE** | **PCE+eGFR** | **PRS** | **PRS+PCE+eGFR** |
| --- | --- | --- | --- | --- | --- | --- |
| **C-index** | 0.7291 | 0.6511 | 0.6612 | 0.6691 | 0.6239 | 0.6536 |
| **PPV 1 yr** | 0.019 | 0.009 | 0.009 | 0.009 | 0.012 | 0.012 |
| **PPV 2 yr** | 0.046 | 0.046 | 0.019 | 0.028 | 0.024 | 0.035 |
| **PPV 3 yr** | 0.102 | 0.074 | 0.037 | 0.056 | 0.047 | 0.059 |
| **PPV 4 yr** | 0.12 | 0.111 | 0.083 | 0.102 | 0.082 | 0.106 |
| **PPV 5 yr** | 0.167 | 0.157 | 0.111 | 0.139 | 0.118 | 0.118 |
| **PPV 6 yr** | 0.185 | 0.157 | 0.111 | 0.139 | 0.118 | 0.118 |
| **PPV 7 yr** | 0.213 | 0.204 | 0.167 | 0.185 | 0.141 | 0.153 |
| **PPV 8 yr** | 0.241 | 0.222 | 0.176 | 0.204 | 0.153 | 0.165 |
| **PPV 9 yr** | 0.278 | 0.231 | 0.185 | 0.231 | 0.165 | 0.2 |
| **PPV 10 yr** | 0.296 | 0.269 | 0.231 | 0.278 | 0.212 | 0.247 |
| **NPV 1 yr** | 0.995 | 0.993 | 0.993 | 0.993 | 0.991 | 0.991 |
| **NPV 2 yr** | 0.984 | 0.984 | 0.977 | 0.979 | 0.976 | 0.979 |
| **NPV 3 yr** | 0.979 | 0.972 | 0.963 | 0.968 | 0.965 | 0.968 |
| **NPV 4 yr** | 0.963 | 0.961 | 0.954 | 0.958 | 0.953 | 0.959 |
| **NPV 5 yr** | 0.956 | 0.954 | 0.942 | 0.949 | 0.944 | 0.944 |
| **NPV 6 yr** | 0.949 | 0.942 | 0.931 | 0.938 | 0.932 | 0.932 |
| **NPV 7 yr** | 0.94 | 0.938 | 0.928 | 0.933 | 0.924 | 0.926 |
| **NPV 8 yr** | 0.94 | 0.935 | 0.924 | 0.931 | 0.921 | 0.924 |
| **NPV 9 yr** | 0.926 | 0.914 | 0.903 | 0.914 | 0.894 | 0.903 |
| **NPV 10 yr** | 0.905 | 0.898 | 0.889 | 0.9 | 0.879 | 0.888 |
